# Development and validation of multivariate predictors of primary endocrine resistance to tamoxifen and aromatase inhibitors in luminal breast cancer reveal drug-specific differences

**DOI:** 10.1101/2023.11.13.23298466

**Authors:** Guokun Zhang, Vindi Jurinovic, Stephan Bartels, Matthias Christgen, Henriette Christgen, Leonie Donata Kandt, Mieke Raap, Janin Klein, Anna-Lena Katzke, Winfried Hofmann, Doris Steinemann, Ronald E. Kates, Oleg Gluz, Monika Graeser, Sherko Kümmel, Ulrike Nitz, Christoph Plass, Ulrich Lehmann, Ulrich Mansmann, Clarissa Gerhäuser, Nadia Harbeck, Hans H. Kreipe

**Author notes:** Corresponding author: Dr. Clarissa Gerhäuser, Division of Cancer Epigenomics, German Cancer Research Center, Im Neuenheimer Feld 280, 69120 Heidelberg, Germany.

## Abstract

**Background:** Endocrine therapy is highly effective in blocking the estrogen receptor pathway in HR+/HER2– early breast cancer (EBC). However, up to 40% of patients experience relapse during or after adjuvant endocrine therapy. Here, we investigate molecular mechanisms associated with primary resistance to endocrine therapy and develop predictive models.

**Patients and Methods:** In the WSG-ADAPT trial (NCT01779206), HR+/HER2-EBC patients underwent pre-operative short-term endocrine therapy (pET). Treatment response was determined by immunohistochemical in-situ labeling of cycling cells (G1 to M-phase) with Ki67 before and after pET. We performed targeted next generation sequencing and Infinium MethylationEPIC-based DNA methylation analysis post-pET in a discovery cohort (n=364, responder (R) and non-responder (NR) pairs matched for clinicopathologic features) and a validation cohort (n=270, unmatched). Predictive indices of endocrine resistance under both treatments were constructed using lasso penalized logistic regression. A subset of breast cancers from ‘The Cancer Genome Atlas’ project (TCGA-BRCA) was used for external validation.

**Results:** TP53 mutations were prominently associated with primary resistance to both tamoxifen (TAM) and aromatase inhibitors (AI), with AI non-responders exhibiting resistance in up to 32% of cases. Additionally, we identified distinct DNA methylation patterns in TAM and AI non-responders, with TAM non-responders showing global DNA methylation loss, associated with KRAS signaling, apical junctions and epithelial-mesenchymal transition (EMT). Conversely, we observed methylation gain in AI non-responders affecting developmental transcription factors, hypoxia and estrogen signaling. TAM or AI resistance was associated with increased methylation-inferred proportions of immune cells and decreased proportions of endothelial cells. Based on these findings and patient age, we developed the Predictive Endocrine ResistanCe Index (PERCI). PERCI stratified NR and R cases in both treatment groups and cohorts with high accuracy (ROC AUC TAM discovery 93.9%, validation 83%; AI discovery 98.6%, validation 76.9%). A simplified PERCI efficiently predicted progression-free survival in the TCGA-BRCA sub-cohort (Kaplan-Meier log-rank p-value = 0.03 between low and high PERCI groups).

**Conclusions:** We identified genomic and epigenomic features associated with primary resistance to TMA and AI. By combining information on genomic alterations, patient age, differential methylation and tumor microenvironment (TME) composition, we developed PERCI TAM and PERCI AI as novel predictors of primary resistance, with potential additional prognostic value. Applying PERCI in a clinical setting may allow patient-specific drug selection to overcome resistance.

WSG-ADAPT, NCT01779206, Registered 2013-01-25, retrospectively registered.

## Background

Endocrine therapy is a highly effective treatment blocking the estrogen receptor pathway in breast cancer (BC). However, up to 40% of patients diagnosed with operable ER and/or PR-positive (*i.e.* luminal) tumors relapse during or after adjuvant endocrine therapy [1]. Clinically, two patterns of resistance to endocrine therapy are recognized: primary, intrinsic resistance and secondary, acquired resistance. Acquired resistance is often attributed to activating mutations in the estrogen receptor (ESR1), whereas the mechanisms of intrinsic resistance are not fully understood [2–4]. In addition, the drug-specific effect of hormone therapy is an unresolved issue. Recent *in vitro* studies suggest that epigenomic mechanisms, including genome-wide reprogramming of the chromatin landscape and DNA methylation changes, contribute to the development of resistance to endocrine therapy [5–8].

Neoadjuvant therapy trials or window trials with short-term endocrine treatment prior to surgery can help to study endocrine resistance. Effective endocrine therapy results in tumor cell growth arrest as evidenced by a decrease in the Ki67 labeling index [9–11]. Persistently high Ki67 expression despite hormone blockade identifies estrogen-independent proliferation, which is associated with an increased risk of disease recurrence and death.

The WSG-ADAPT trial, which enrolled to date more than 5600 patients with luminal BC, 2290 of whom in the endocrine-therapy subtrial, provides a novel approach to investigate endocrine resistance [12]. More than 70% of patients in the ADAPT trial responded to short-term endocrine therapy with either tamoxifen or aromatase inhibitors. Non-responding tumors maintain their original growth rate despite hormone blockade, making them ideal candidates to study the mechanisms of primary endocrine resistance.

In this study, a discovery and validation cohort of endocrine therapy responsive and non-responsive patients was sub-sampled from the WSG-ADAPT trial (summarized in **Fig. 1**). We aimed to identify molecular mechanisms associated with the resistant phenotype and common pathways recurrently affected by different aberrations using targeted next-generation sequencing and Infinium MethylationEPIC BeadChip-based DNA methylation analysis. We hypothesized that knowledge of the genetic and epigenetic aberrations in BC that are associated with endocrine resistance could provide biomarkers for the aggressive subtype of hormone receptor-positive luminal breast cancer and allow the development of predictive models for the response to endocrine therapy. As an external validation cohort, we planned to use a subset of the TCGA-BRCA cohort that was clinically matched to patients in our cohorts. The results of this study provide insight into the mechanisms of resistance to tamoxifen and aromatase inhibitors, and thus suggest a way to overcome resistance by switching to the drug that is not affected.

**Fig. 1:**
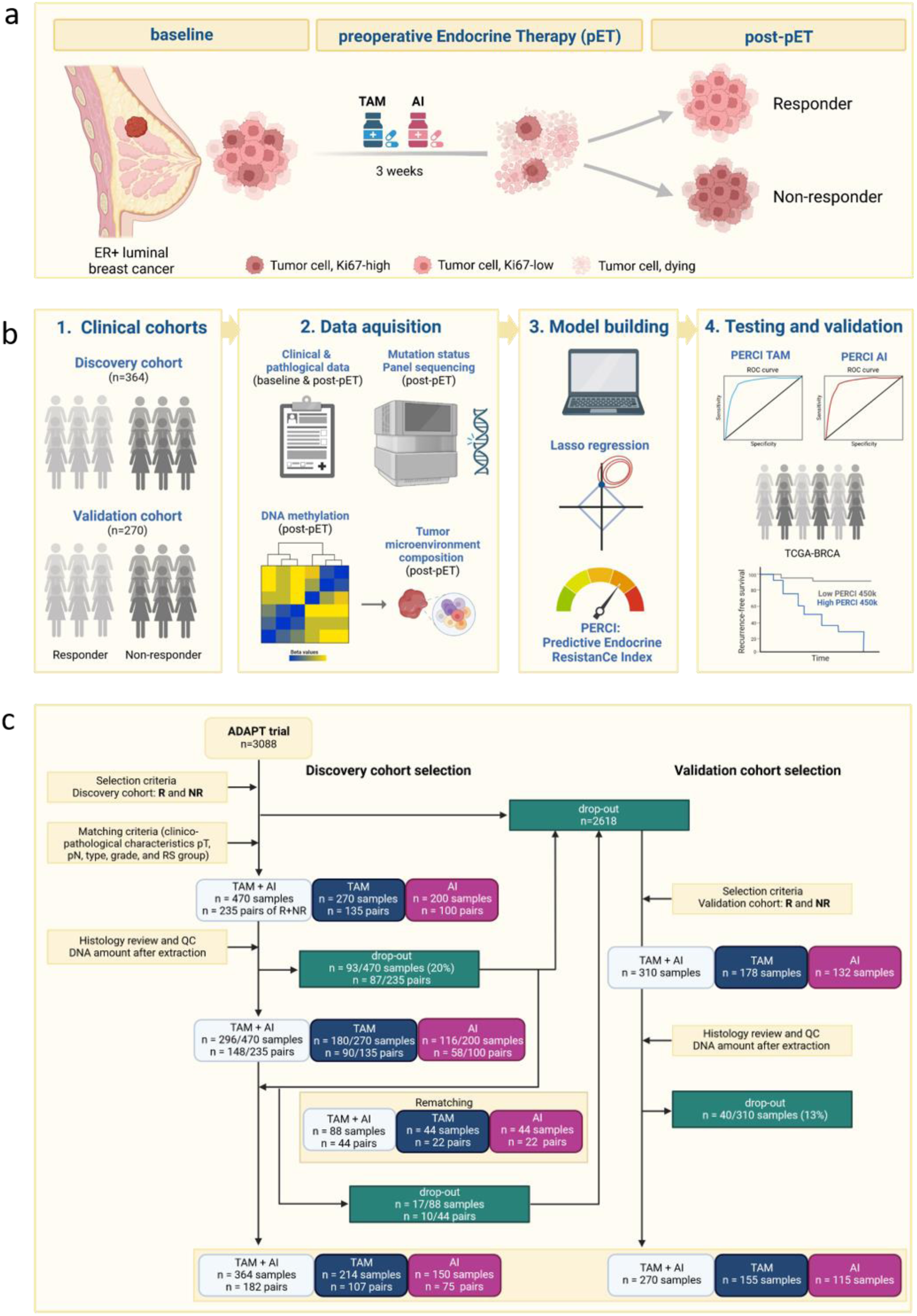
Concept of the study. (a) In the WSG-ADAPT trial, patients with ER+ luminal BC were randomized to receive three weeks of preoperative endocrine therapy (pET) with tamoxifen (TAM) or aromatase inhibitors (AI). Treatment response was estimated by Ki67 staining post-pET compared to baseline staining. (b) In this study, we aimed to develop models to predict response to endocrine therapy. We selected patients for a discovery and a validation cohort and collected clinico-pathologic data, mutation status of selected cancer driver genes, and DNA methylation data at baseline and/or post-pET. For external validation, we used a subset of the TCGA-BRCA cohort that matched the selection criteria for our cohorts. (c) Flowchart of the sample selection for the discovery cohort (left, matched sample design) and the validation cohort (right, un-matched design). The numbers reflect the case selection for molecular examinations as of the beginning of the study. At this time, patients were still being recruited in ADAPT. Figure designed using BioRender.

## Methods

### Study design and sample collection

The ADAPT trial (NCT01779206 [12,13]) was a phase II, multicenter, controlled, nonblinded, randomly assigned, investigator-initiated trial performed in the framework of WSG-ADAPT umbrella protocol and at the Institute of Pathology, Hannover Medical School (MHH). We selected patients with early breast cancer who had ER-positive and/or progesterone receptor (PR)-positive, human epidermal growth factor receptor 2 (HER2)-negative tumors. The patients had received three weeks of preoperative endocrine therapy (pET) (tamoxifen (TAM) in premenopausal women and aromatase inhibitor (AI) in postmenopausal women). Baseline and post-pET tumor biopsies were obtained from all patients, and formalin-fixed, paraffin-embedded (FFPE) specimens were prepared. The Oncotype DX Recurrence Score (RS) of the baseline samples was determined at the Genomic Health Inc. laboratory, and all baseline and post-pET FFPE samples were submitted to immunohistochemistry at the ADAPT Study Central Reference pathology at Hannover Medical School (MHH). The expression of ER, PR, HER2, and the Ki67 cell proliferation marker were assessed using standardized methods as previously described [12]. Tumors with baseline Ki67 < 35% and of PR > 20% were classified as luminal A, whereas tumors with baseline Ki67 ≥ 35% and PR ≤ 20% were designated as luminal B subtypes.

Response to endocrine therapy was determined by post-pET Ki67 and Ki67 decrease from baseline. In the discovery cohort (n=364) patients with post-pET Ki67 <10% and a relative decrease ≥ 70% were considered responders (R), and those with post-pET Ki67 ≥ 20% and a relative decrease ≤ 20% were considered non-responders (NR). The discovery cohort was divided into TAM (n=214) and AI treatment groups (n= 150), each with R and NR patients matched for histopathologic features, pT, pN, ER, PR, HER2 status and RS score at baseline. Tumors (R/NR) were also balanced for WSG central histologic grade. We included all sample pairs that met our selection criteria (flowchart in **Fig. 1c**, **Supp. Table S1a**). For the validation cohort (n= 270), we relaxed the selection criteria to allow inclusion of additional patients. Responders were defined as post-pET Ki67 <10%, and non-reponders had post-pET Ki67 ≥ 20%. The response groups were not matched with respect to histopathologic features, RS score, or histologic grade. We included 75 TAM responders and 80 non-responders, and 67 AI responders and 48 non-responders **(Fig. 1c, Supp. Table S1b)**. The TCGA-BRCA sub-cohort (n = 269) was sub-sampled from the TCGA-BRCA cohort to provide external validation of the prediction models. Selection criteria included *ER status positive*, *HER2 status non-positive*, *distant metastasis free (M0)*, *no prior neoadjuvant therapy*, and *progression-free survival (PFS) information available* (**Supp. Table S2**). Samples were divided into two groups based on menopausal status: TAM-like for premenopausal patients (n = 75) and AI-like for postmenopausal patients (n = 194). Histologic grade was determined using WHO criteria (information on tubule formation, pleomorphism, and cell proliferation from [14]).

### Statistical analyses

In the discovery cohort, McNemar’s test for symmetry was used to compare R and NR patients for clinical variables. In the validation cohort, the chi-squared test for trends was used to compare *Ki67 at baseline* and *Ki67 post-pET*, and Fisher’s exact test was used for all other comparisons. The relationship between clinical variables and TAM and AI prediction scores was calculated using Spearman’s rank correlation coefficient.

### Next generation target sequencing and variant calling

After manual microdissection to enrich for invasive tumor cells, DNA was extracted from FFPE tissue using the Maxwell RSC DNA FFPE kit on a Maxwell RSC instrument (Promega). Quality control tests were performed to ensure that the DNA samples (n=362 from the discovery cohort and n=222 from the validation cohort) met the required criteria. Next-generation sequencing (NGS) was performed at the MHH using a S5 prime instrument (ThermoFisher Scientific). Two NGS panels were used to cover the complete protein coding regions of known driver genes and genes associated with endocrine resistance, as described previously [15]. The first panel was the Oncomine Comprehensive v3 assay (ThermoFisher Scientific), which included 161 genes and had the ability to also detect copy number variations. The second panel was a custom-made panel including the full-coding sequence of 17 genes (*ABCA13, CBFB, CDH1, ERBB2, ERCC2, ESR1, FAT1, FAT2, FAT3, GATA3, MAP3K1, MUTYH, PIK3CA, RUNX1, RYR2, TBX3, TP53*) to add regions of genes which are frequently mutated in BC and missing in the Oncomine Comprehensive v3 assay. Variant calling and functional annotation were performed using ANNOVAR software (http://annovar.openbioinformatics.org/en/latest/). Gene deletions were selected using a copy number cutoff ≤ 0.6 (upper CI ≤ 0.75). Gene amplifications were selected using a copy number cutoff ≥ 5.0 (lower CI ≥ 3.5). The results were manually curated to exclude FFPE-related genetic artifacts. Statistical analysis was performed using Fisher’s exact test to compare frequencies of recurrent genomic alterations (RGA) between responders and non-responders in each treatment group. Only genes with a RGA frequency of at least 7.5% in one of the subgroups (TAM, AI, NR, R in the discovery or validation cohorts) were included in the analysis.

### DNA methylation pre-processing and cell type deconvolution

DNA (range 84-1400 ng) was submitted to the DKFZ Genome and Proteome core facility for DNA methylation analysis using HumanMethylationEPIC (EPIC) BeadChips (Illumina, CA, USA), including an additional restore step for FFPE material. We downloaded reference datasets from GEO, including data for six immune cell types from six healthy subjects (GSE110554), human mammary fibroblast, human mammary epithelial cells, human mammary endothelial cells (GSE74877) [16], and human breast cancer cells (MCF7, GSE68379)[17]. Raw data were preprocessed using the RnBeads R package version 2.15.1 [18,19], with beta-mixture quantile normalization and no background correction. We filtered out probes with a SNP overlapping with the C nucleotide of the CpG site and a MAF > 0.01 (dbSNP 150), as well as probes with the last 3 bases in their target sequence overlap with a SNP (MAF > 0.05). We also removed cross-hybridizing probes [20,21] and non-CpG probes. We estimated the cell type composition using the Houseman algorithm [22] implemented in RnBeads, using the settings inference.max.cell.type.markers = 100000, inference.top.cell.type.markers = 500, and adjusted the results proportionally to a sum of 100%. For differential methylation analyses, raw data were preprocessed using the SeSAMe package [23] with linear dye-bias correction (dyeBiasCorr) and background subtraction using “noobsb”. We performed differential methylation analyses using “limma”, with age and sample processing (bulk or macrodissection) used as covariables. We defined differentially methylated CpG sites (DMS) as having a methylation difference > 10% and p-value < 0.005. To annotate DMS to genes, we developed a pipeline to map CpG sites represented on the EPIC array to the next transcription start site (TSS) using hg19 transcript information from the R package “EnsDb.Hsapiens.v75” [24]. The pipeline is available on Github at https://github.com/gk-zhang/InfiniumEPICMethylation.hg19/tree/main. We performed gene set overrepresentation analyses using Molecular Signatures Database (MSigDB) hallmark gene sets [25]. DMS were annotated with chromatin states using published ChromHMM classification for MCF7 cells [26]. A consensus list of partially methylated domains (PMDs) in human breast cancer was constructed from Brinkman et al. [27], with occurrence in ≥9 of 30 tumors and a size > 100kb (n= 2538 regions). We calculated MeTIL scores as described in [28]. For the TCGA-BRCA cohort, we downloaded Illumina HumanMethylation450 data from GDC (Genomic Data Commons Data Portal, https://portal.gdc.cancer.gov) and preprocessed the data with RnBeads using SeSAMe-adapted settings with *scaling.internal* for normalization and background subtraction using *noobsb*. We selected CpG sites overlapping between the EPIC and the 450k datasets and further preprocessed the data as described above. We built the 450k models using the overlapping CpG sites.

The gene regulatory potential of our DMS was tested in the TCGA-BRCA sub-cohort matching our samples (n = 269) and DMS covered on the 450k array. Gene expression HTSeq FPKM-UQ quantification data were downloaded with the function GDCquery of R package TCGAbiolinks (version: 2.22.1) and used to compute Pearson and Spearman correlations between DMS beta values and log2 expression values.

### Building of the ‘Predictive Endocrine ResistanCe Index’ PERCI

Predictive indices of endocrine resistance under either treatment were constructed within both discovery cohorts using lasso penalized logistic regression with the R package “glmnet” (version 4.1-3). Internal cross-validation was used to determine model hyperparameters [29] based on optimizing the quality of out-of-sample-prediction in terms of discrimination by ROC-AUC [30]. Only the samples with complete data for all characteristics were included (as shown in **Supplementary Table S3**). For each treatment group, the classifier was built using the beta values of the DMS with a minimum mean methylation difference of 10% between the responder and non-responder groups. In addition, we used the RGA (set as 0 for wild-type and 1 for detected alterations) with significantly different alteration frequencies (*p<0.025*) between the response groups. Other candidate predictor variables included patient age and cell type information, which was obtained from the methylation data. The prediction outcome was defined as the endocrine resistance outcome with a binary classification of R and NR. Different prediction models were developed by applying different cut-off p-values for differential methylation for the TAM and AI groups, respectively. The performance of the prediction models was evaluated by calculating the area under the receiver-operating curve (AUC) between the known classification result and the prediction scores (described below) by using the R package pROC (version: 1.18.0). The final models were selected based on the best AUCs in the validation cohort data of the TAM and AI groups, respectively.

Each model is composed of predictors and coefficients. PERCI TAM and AI scores were calculated as the weighted sum of the predictor values multiplied by the model coefficients. The prediction scores were scaled to the range [0, 1] by dividing the original score by the difference between the theoretical maximum score and the theoretical minimum score. The theoretical maximum/minimum score was obtained by assigning a (beta) value of 1/0 to CpG sites or mutated genes with positive coefficients and 0/1 to those with negative coefficients. For patient age and cell type information, the median values of the discovery cohort data were used for the calculation of both theoretical maximum and minimum score.

*Accuracy*, *precision (= positive predictive value PPV)* and *recall* from the output of the R package pROC were used as metrics to evaluate the performance of the prediction models. As a cutoff we defined the scaled PERCI score with the highest accuracy. *F1 score* was calculated as *2×precision×recall/(precision+recall)*. In the confusion matrix, responders are defined as controls and non-responders as cases.

### Constructing PERCI 450k

To ensure wider applicability to publicly available Illumina 450k methylation datasets, we developed additional classifiers specifically tailored to this platform, limiting the input to CpG sites covered on the 450k array. We included methylation data from CpG sites with a minimum mean methylation difference of 5% between the responder and non-responder groups and p-values for differential methylation of *p ≤ 0.05 to p ≤ 0.00005*. We also added patient age as a variable to the model. The same approach as described above was used to construct PERCI 450k models and produce the scaled scores.

Performance of the PERCI 450k models was tested in patients from the TCGA-BRCA sub-cohort, subdivided in TAM-and AI-like groups. As endocrine treatment and response information was not available, the models were evaluated based on their ability to predict tumor progression-free survival (PFS) using the prediction scores. Survival analyses were performed using the R package “survival” (version 3.5-0), using the information of time to event (progression or censoring), limited to a follow-up time of 120 months. PERCI TAM 450k and PERCI AI 450k scores were used to classify the patients into *low* and *high* groups. The cutoffs were chosen by minimizing the p-value of the log-rank test. Fisher’s exact test was used to test the significant difference of clinic-pathological variables between the PERCI 450k score high and low groups (**Supp. Table S2**).

## Results

### Histologic grade changes correlate with Ki67 staining in response to preoperative endocrine therapy (pET)

The response groups in the discovery cohort were well matched for clinico-pathologic characteristics at baseline (**Suppl. Table S1a, S3**). Cases treated with TAM tended to be of lower grade than those treated with AI. Response to short-term pET was reflected by changes in histologic grade from baseline to post-pET (**Fig. 2a**). In both treatment groups, >40% of responders decreased in grade, while many non-responders had a higher grade after pET (**Supp. Fig. S1a**). For both the TAM and AI groups, Spearman’s rank correlation analysis confirmed high correlation between histologic grade and Ki67 staining at baseline and after pET (TAM baseline ρ=0.54, TAM post-pET ρ=0.69; AI baseline ρ=0.61, AI post-pET ρ=0.79, fdr-corrected p-values *< 0.01*) (**Supp. Fig. S2a**). In the AI group, we observed a significant reduction in PR staining after pET compared to baseline (AI responders *p = 2.54e-08*, AI non-responders *p=5.32e-04*) (**Fig. 2c**). Combining Ki67 and PR staining information, we divided the patients into luminal A and B subtypes (**Supp. Table S1a, S3**). Over 40% of AI cases, but only 10% (R) and 19% (NR) of the TAM cases were categorized as LumB (**Fig. 2d**). Similar differences were also reflected in the OncotypeDX RS group distribution. Patients in the TAM group were predominantly in RS group 1 (28%) and RS group 2 (64%), whereas 48% of the AI-treated cases were in RS group 2 and 40% in RS group 3 (**Fig. 2e**). Consequently, we observed a positive correlation with luminal subtypes (Kendall rank correlation test, TAM τ = 0.33; AI τ = 0.51, fdr-corrected p-value < 0.01)(**Supp. Fig. S2c**). In the TAM group, RS group annotations were weakly negatively correlated with hormone receptor staining at baseline (ER, ρ = –0.31; PR, ρ = –0.33, fdr-corrected p-values *< 0.01*). For the AI-treated cases, anti-correlation of RS score and PR staining was even more pronounced (baseline, ρ = –0.46; post-pET ρ = –0.47, fdr-corrected p-values *< 0.01*)(**Supp. Fig. S2a**).

**Fig. 2:**
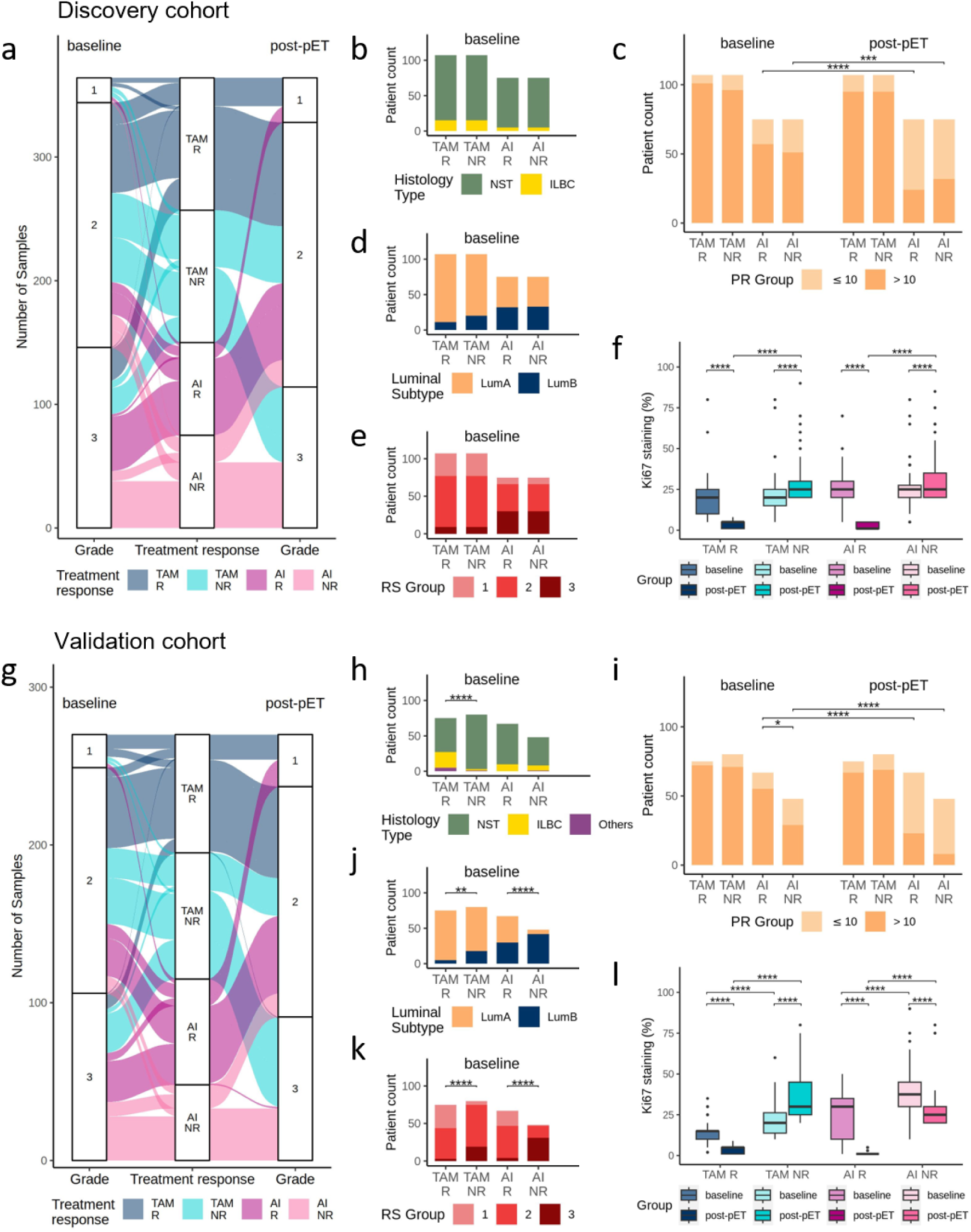
Descriptive statistics of the cohorts. (a – f) Description of the discovery cohort (n = 364, TAM = 214, AI = 150), (g – l) description of the validation cohort (n = 270, TAM = 155, AI = 115). Distribution of patients (R = responder, NR = non-responder) before (baseline) and after anti-hormone (post-pET) treatment according to: (a, g) tumor grade; (b, h) histology type; (c, i) progesterone receptor (PR) status; (d, j) luminal subtype; (f, l) percentage of Ki67 positive staining in IHC. (e, k) Distribution of patients before endocrine therapy according to recurrence score (RS). Statistical differences of numerical variables between matched pairs as well as for baseline and post-pET comparisons were tested using paired Wilcoxon tests. All other comparisons of numerical variables were analyzed using non-paired Wilcoxon tests. Statistical differences of categorical variables between matched pairs as well as for baseline and post-pET comparisons were analyzed using McNemar test. All other comparisons of categorical variables were analyzed using Fisher’s exact test. For all statistical tests, asterisks *, **, ***,**** indicate p-values < 0.05, 0.01, 0.001, 0.0001.

As no matching protocol was used to select cases for the validation cohort, significant differences between responders and non-responders were observed in both treatment groups (TAM: histology, pT, grade, baseline Ki67, luminal subtype, RS group; AI: grade, baseline PR and Ki67 staining, luminal subtype, RS group) (**Supp. Table S1b, S3**). pET responders had consistently lower grades at baseline than non-responders in both treatment groups (TAM *p=7.48e-04*, AI *p=1.67e-03*)(**Fig. 2g**). Again, changes in histologic grading from baseline to post-pET reflected response or resistance to pET (**Supp. Fig S1b**). Grade and Ki67 staining were highly correlated at baseline and post-pET in both treatment groups (TAM baseline ρ=0.67, TAM post-pET ρ=0.74; AI baseline ρ=0.70, AI post-pET ρ=0.77, fdr-corrected p-values *< 0.01*) (**Supp. Fig. S2b**), confirming our findings in the discovery cohort. We observed a significantly higher proportion of invasive lobular breast cancer (ILBC) in TAM responders than in non-responders (R 29%, NR 3%), but not in AI-treated cases, with 15% ILBC in both response groups (**Fig. 2h**). Due to the unpaired selection protocol in the validation cohort, significantly (*p=0.015*) more AI non-responder than responders had weak PR staining (PR ≤ 10%) prior to pET. We confirmed the decrease in PR staining after AI treatment in both response groups (AI-R *p=1.06e-07*, AI-NR *p=6.33e-05*) (**Fig. 2i**). In both treatment groups, non-responders were significantly more often annotated as LumB subtype (Fisher’s exact test TAM *p=6.2e-03, AI = 2.1e-06*) (**Fig. 2j**) and had higher baseline RS scores than responders (Fisher’s exact test TAM *p=1.9e-08*, AI *p= 2.1e-12*) (**Fig. 2k**, and **Supp. Table S1b**). RS scores of TAM-treated cases were positively correlated with grade at baseline and post-pET (ρ= 0.41 and 0.41, fdr-corrected p-values *< 0.01*), and the latter observation was even stronger in AI-treated cases (ρ= 0.56, fdr-corrected p-value *< 0.01*). In addition, RS scores in AI-treated cases showed a strong negative correlation with baseline PR staining (ρ = –0.5, fdr-corrected p-value *< 0.01*) (**Supp. Fig. S2b**). Finally, in both treatment groups, Ki67 staining at baseline was higher in non-responders than in responders (TAM *p=1.18e-07*, AI *p=2.05e-05*) (**Fig. 2l, Supp. Tables S1b**).

In summary, TAM cases had lower grade and RS scores than AI cases in both cohorts. pET resulted in consistent changes in grade in both treatment groups, with responders lowering grade and non-responders changing to a higher grade. Grade was highly correlated with Ki67 staining at both baseline and post-pET. PR staining was consistently lower post-pET than at baseline in AI responders and non-responders in both cohorts.

### Recurrent TP53 mutations in luminal BC promote resistance to pET

We performed NGS panel sequencing to identify recurrent genomic alterations (RGA) that might indicate pET resistance (**Supp. Table S4**). Mutations of *PIK3CA*, *GATA3*, *MAP3K1*, and amplifications of *CCND1*, *FGF3* and *FGF19* on chr11q13.3 were most frequently detected in both cohorts (**Fig. 3a,b**), confirming previously published observations [31–34]. None of these RGA were found to be significantly altered between R and NR cases in either TAM or AI treatment groups. The tumor/invasion suppressor gene E-cadherin (*CDH1*) is mutated in the majority of ILBC cases [33,35]. We detected frameshift and splice-site mutations in 10% and 16% of the patients in the discovery and validation cohort, respectively, and confirmed significant enrichment of *CDH1* mutations in ILBC (discovery cohort p=*8.5e-11*, validation cohort p<*2.2e-16*), with strong positive correlation with histology type (Kendall rank correlation τ = 0.61 to 0.81, fdr-corrected p-value *< 0.01,* **Supp.** Fig. 2c,d). *CDH1* mutations were strongly anti-correlated with E-cadherin protein expression at baseline and post-pET in both treatment groups and cohorts (τ = –0.59 to –0.91, fdr-corrected p-value *< 0.01,* **Supp.** Fig. 2c,d).

**Fig. 3.**
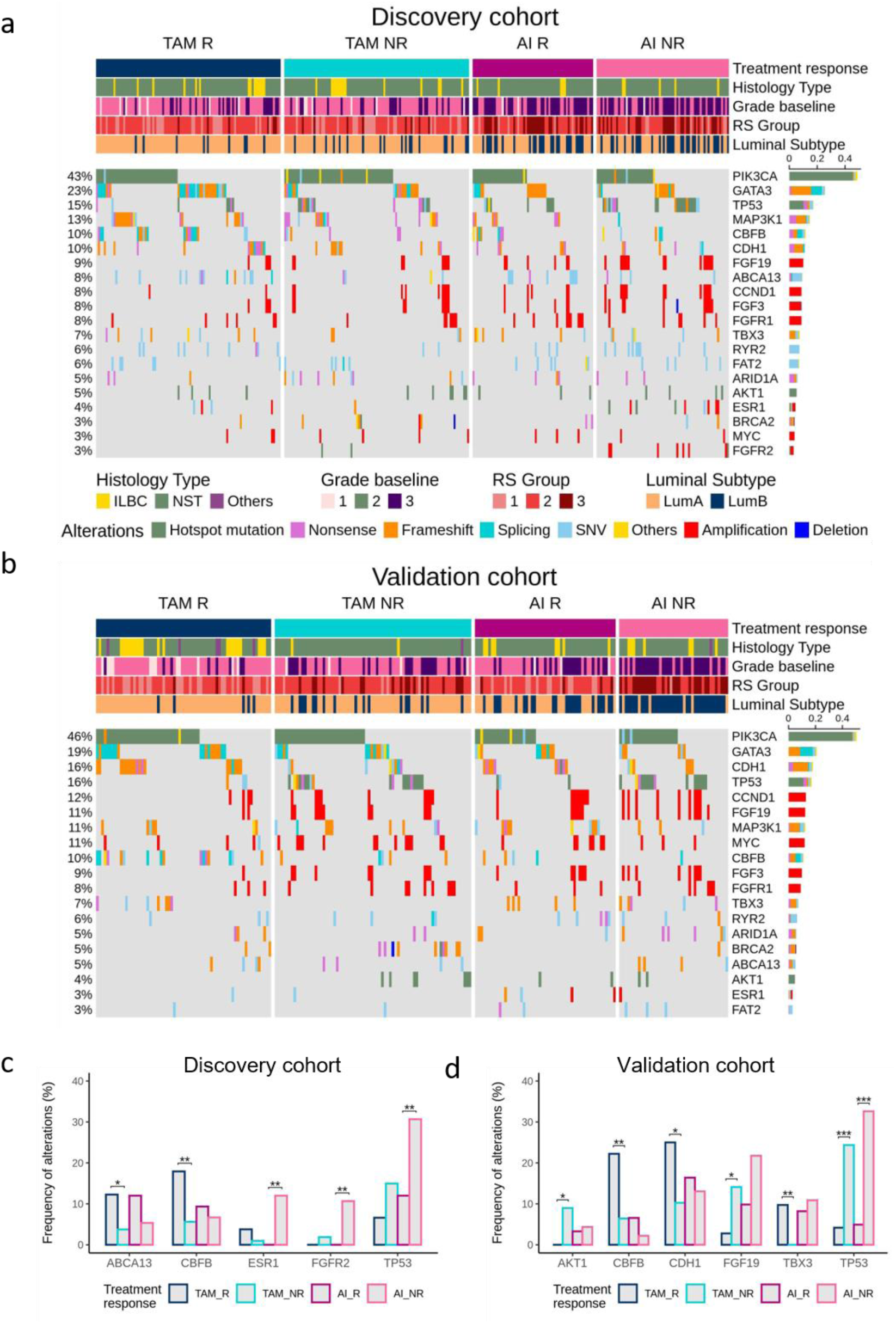
Recurrent genomic alterations (RGA) in the (a) discovery cohort, and (b) validation cohort. Legend for (b) as in (a). Mutations, amplifications and deletions in post-pET samples, sorted by total alteration burden per cohort. Only RGA at minimum 7.5% recurrence in either sub-group are shown. The oncoprints summarize the mutational landscape of RGA regions in the present study, color-coded by the mutational event-type and separated into TAM R, TAM NR, AI R and AI NR. Clinical annotations of cases are indicated on the top. The barplot at the right quantifies the recurrence of each RGA in each cohort. (c, d) Frequencies of RGA with significant differences in discovery (c) and validation (d) cohort, statistical analyses using Fisher-exact test with *, **, *** p-value < 0.025, 0.01, 0.001.

In the discovery cohort, we detected point mutations in the ATP-binding cassette transporter 13 (*ABCA13*) gene significantly more often in the TAM R group than in the TAM NR group (12.2% vs. 3.7%, *p=0.024*) (**Fig. 3c**). *CBFB* mutations were also significantly enriched in the TAM R group (17.7% vs. 5.6%, *p=0.006*) (**Fig. 3c**). This finding was confirmed in the validation cohort (TAM R 22.2%, TAM NR 6.4%, *p=0.008*) (**Fig. 3d**) and positively correlated with *GATA3* alterations (Kendall rank correlation, discovery cohort τ=0.3, validation cohort τ=0.3, fdr-corrected p-value *< 0.01*) (**Supp. Fig. S2c**).

In the AI discovery cohort, but not the validation cohort, we observed significantly more *ESR1* alterations in non-responders than in responders (12% vs. 0%, *p=0.003*) (**Fig. 3c**). Also, *FGFR2* amplifications were detected more frequently in the AI-NR group than in the AI-R group (9.3% vs. 0%, *p=0.014*) (**Fig. 3c**) and were positively correlated with histologic grade and Ki67 status (τ=0.29 and τ=0.27, fdr-corrected p-values *<0.01*) after treatment, as well as with *TP53* mutations (τ=0.31, fdr-corrected p-value *<0.01*)(**Supp. Fig. S2c**). As recently described by our group, *TP53* mutations are significantly associated with therapy resistance to pET in both TAM-and AI-treated cases [15,36]. Consistent with these reports, significant enrichment of *TP53* mutations in non-responding patients of both treatment groups in the discovery cohort (TAM *p=0.032*, AI *p=0.011*)(**Fig. 3c**) was confirmed in the validation cohort (TAM *p=0.0008*, *fdr=0.099*; AI *p=0.00015, fdr=0.019*)(**Fig. 3d**).

In the validation cohort, TAM non-responders harbored significantly more alterations in AKT1 (NR 9.0% vs. R 0%, *p=0.014*) and amplifications of *FGF19* (NR 12.8% vs. R 2.8%, *p=0.033*) than TAM responders (**Fig. 3d**). Conversely, we detected significantly more *TBX3* mutations in TAM responders (9.7%) than in non-responders (0%, *p=0.005*) (**Fig. 3d**). These significant differences were not observed in the discovery cohort. In TAM-treated cases, amplifications of *CCND1*, *FGF3*, and *FGF19* on chr11q13.3 were positively correlated with grade (τ= 0.28 to 0.34, fdr-corrected p-values *< 0.01*), Luminal Subtype (τ= 0.27 to 0.34, fdr-corrected p-values *< 0.01*) and baseline Ki67 (τ= 0.32 to 0.36, fdr-corrected p-values *< 0.01*) (**Supp. Fig. S2d**).

To summarize, we found several significant differences in the incidence of RGA between responding and non-responding cases. TAM responders had consistently more *CBFB* mutations than non-responders in both cohorts. Amplifications of *ESR1* and *FGFR2* were selective for the AI NR group. With a frequency of up to 32% in AI NR cases, *TP53* mutations were most frequently associated with the development of resistance to endocrine therapy with TAM and AI.

### Different alterations in the DNA methylome and tumor microenvironment contribute to resistance to pET with TAM and AI

Previous studies have reported that endocrine therapy resistance in breast cancer cell lines leads to adaptations in chromatin structure and DNA methylome [5–7]. In addition to our mutation screen, we performed methylation analyses on tumor tissue obtained post-pET using EPIC arrays covering 850.000 CpG sites. In the discovery cohort, we detected 472 significantly (p*<0.005*) differentially methylated CpG sites (DMS) with ≥ 10% mean methylation difference between TAM NR and R groups, and 435 DMS between AI NR and R (**Supp. Fig. S3a**, DMS highlighted in red, **Supp. Table S5**). Nearly 70% of the TAM-DMS and 40% of the AI-DMS were also differentially methylated in the validation cohort (**Supp. Fig. S3b**).

Notably, we observed distinct methylation changes in the two treatment groups. In TAM-treated cases, 90% of DMS were less methylated in non-responders compared to responders (**Fig. 4a, NR<R indicated in dark blue on the right**). Global loss of methylation has been associated with accelerated cell proliferation and the inability of a cell to re-methylate DNA in late-replicating regions after DNA doubling. This leads to the formation of ‘partially methylated domains’ (PMDs) in the nuclear periphery [37]. Using consensus PMD information from a recent breast cancer study [27], we confirmed that nearly 70% of all TAM DMS are located in PMDs (**Fig. 4a, shown in dark green on the right**). We further annotated the DMS with chromatin regions using information from the ER-positive MCF7 cell line [26]. We found that 80% of TAM DMS were located in tightly packed heterochromatin and repressed regions, which overlapped strongly with PMD regions (**Fig. 4a, Supp. Fig. S3c, upper panel**).

**Fig. 4.**
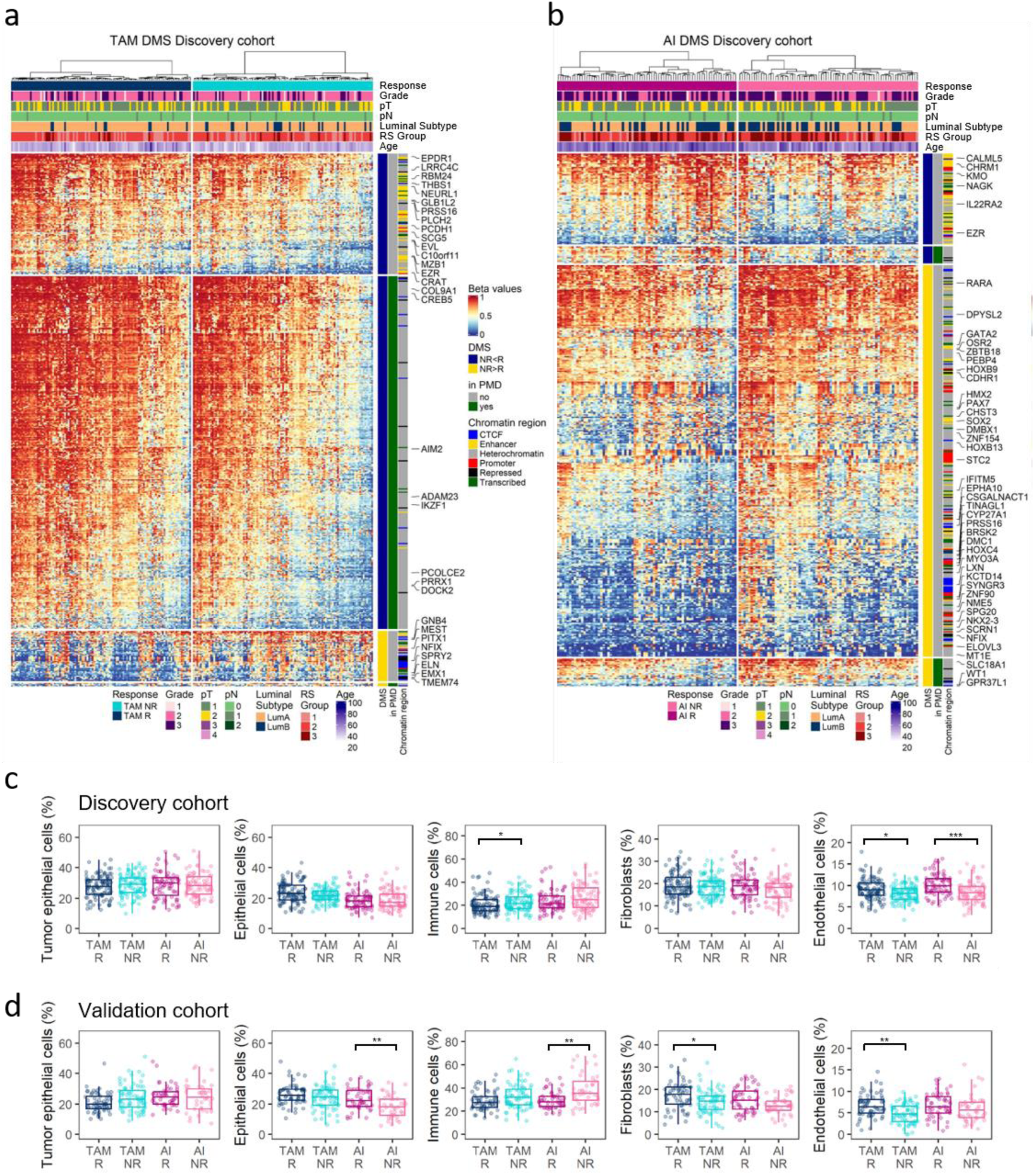
pET-specific alterations in the methylome and tumor microenvironment. (a,b) Heatmaps of methylation beta-values of significant differentially methylated CpG sites (DMS) for TAM-(a) and AI-treated cases (b). Clinical annotations of cases are indicated at the top. DMS with gain (NR>R) and loss (NR<R) in methylation in the non-responder group, breast cancer partially methylated domains (PMDs), localization of DMS in MCF7-derived chromatin regions, and gene symbols of selected genes are indicated on the right. Rows (CpG sites) and columns (cases) are clustered by Euclidean distance and ward.D linkage. The heatmaps are split by response groups (columns) and by methylation change and location in PMDs (rows). Legend for heatmap and side annotations in (b) as in (a). (c,d) Cell type composition of breast cancer samples analyzed using a reference-based deconvolution method in the discovery (c) and validation cohort (d). R and NR groups per pET were compared using Wilcoxon test with *, **, *** fdr-adjusted p-values < *0.05, 0.01, 0.001*.

In contrast to the TAM group, approximately 80% of DMS were higher methylated in AI NR vs. AI R cases (**Fig. 4b, NR>R shown in yellow on the right**). The majority of AI DMS were located outside of PMDs, suggesting a gene-regulatory function. Indeed, about 40% of the DMS were located in enhancer and promoter regions or overlapped with insulator protein CTCF binding or transcribed regions (**Supp. Fig. S3c**, lower panel). Gene set overrepresentation analysis using MSigDB hallmark gene sets suggested that resistance to TAM pET may be associated with KRAS signaling, apical cell-cell junctions and epithelial-mesenchymal transition (**Supp.** Fig. 3d).

We used data for the TCGA-BRCA sub-cohort as a surrogate to calculate correlation coefficients for correlations between methylation and gene expression for the subset of DMS represented on Illumina 450k arrays (**Supp. Table S5**). Although most of the AI DMS had higher methylation levels in NR vs. R, indicating gene silencing, we identified a group of developmental transcription factors with positive correlations between methylation and gene expression, indicating gene upregulation. In addition, AI-DMS-associated genes were enriched in gene sets related to hypoxia and estrogen response (**Supp.** Fig. 3d).

Several lines of evidence suggest that the tumor microenvironment (TME) is involved in the development of resistance to TMA and AI treatment [38]. We used publicly available reference methylation datasets for various cell types (Methods) to estimate the composition of the TME in our samples. In pET-resistant patients, we consistently computed higher immune cell proportions, accompanied by reduced proportions of fibroblasts and endothelial cells (**Fig. 4c,d**). The percentage of stromal tumor-infiltrating lymphocytes in pathologic tissue sections (PaTILs) significantly correlated with TIL levels derived from methylation data (MeTILs) (discovery cohort, ρ = 0.555, *p < 2.2e-16*; validation cohort ρ = 0.464, *p = 3.2e-13,* **Supp. Fig. S3e**). Our MeTIL percentages also highly correlated with MeTIL scores calculated according to [28] (discovery cohort, ρ = 0.638, *p < 2.2e-16*; validation cohort ρ = 0.697, *p < 2.2e-16*). When stratified according to immune cell type, CD4+ cells were most abundant, but increased immune cell infiltration in pET-resistant tumors was not linked to any particular immune cell type (**Supp. Fig. S3f,g**).

Overall, differential methylation analysis revealed distinct profiles of methylation changes in breast cancer patients treated with TAM and those treated with AI. TAM DMS were often less methylated in the NR vs. R group and were associated with PMDs. Conversely, AI DMS were more highly methylated in the NR vs. R group, located in gene regulatory regions and associated with developmental transcription factors, hypoxia and estrogen signaling. In both treatment groups, we computed changes in TME cell composition with increased immune cell infiltration and decreased fibroblast and endothelial cell proportions associated with treatment resistance.

### Developing the ‘Predictive Endocrine ResistanCe Index’ PERCI

After identifying significant differences between responders and non-responders in the discovery cohort, we used lasso penalized logistic regression on these features to train classifiers for TAM and AI resistance, which we named ‘Predictive Endocrine ResistanCe Index’ (PERCI) (**Fig. 5a**). PERCI TAM consists of age information, endothelial cell content, ABCA13 mutations, and methylation data for 29 TAM DMS with different weights (**Fig. 5b, left panel**). Most of the predictive CpG sites were hypomethylated in the non-responder groups, and about half of them were located in heterochromatic regions (**Supp. Tables S3, S5**). Five selected CpG sites (cg15042080, cg16766325, cg04286030, cg18396984, cg15332750) had higher methylation in non-responders than in responders. The inverse correlation of cg16766325 methylation, located in the *SPRY2* promoter, with gene expression in the TCGA-BRCA sub-cohort (Pearson correlation r = –0.423, fdr-adjusted p-value *= 4.11E-12*) may indicate functional relevance in gene regulation. The individual predictors achieved an area under the receiver operating characteristic curve (ROC AUC) of 54.8 (for ABCA13 mutations) to 72% (for cg01838965) in the discovery cohort (**Supp. Fig. S4**). By combining all features, PERCI TAM stratified responders and non-responders with a ROC AUC of 93.9%, (**Fig. 5c**, left panel), with very good accuracy and positive predictive value (PPV) (**Table 1**). Strong performance was confirmed in the validation cohort, with a ROC AUC of 83%. Testing frequencies of correctly and incorrectly annotated cases in a confusion matrix suggests that the PERCI TAM is better at predicting NR than R in both cohorts.

**Fig. 5.**
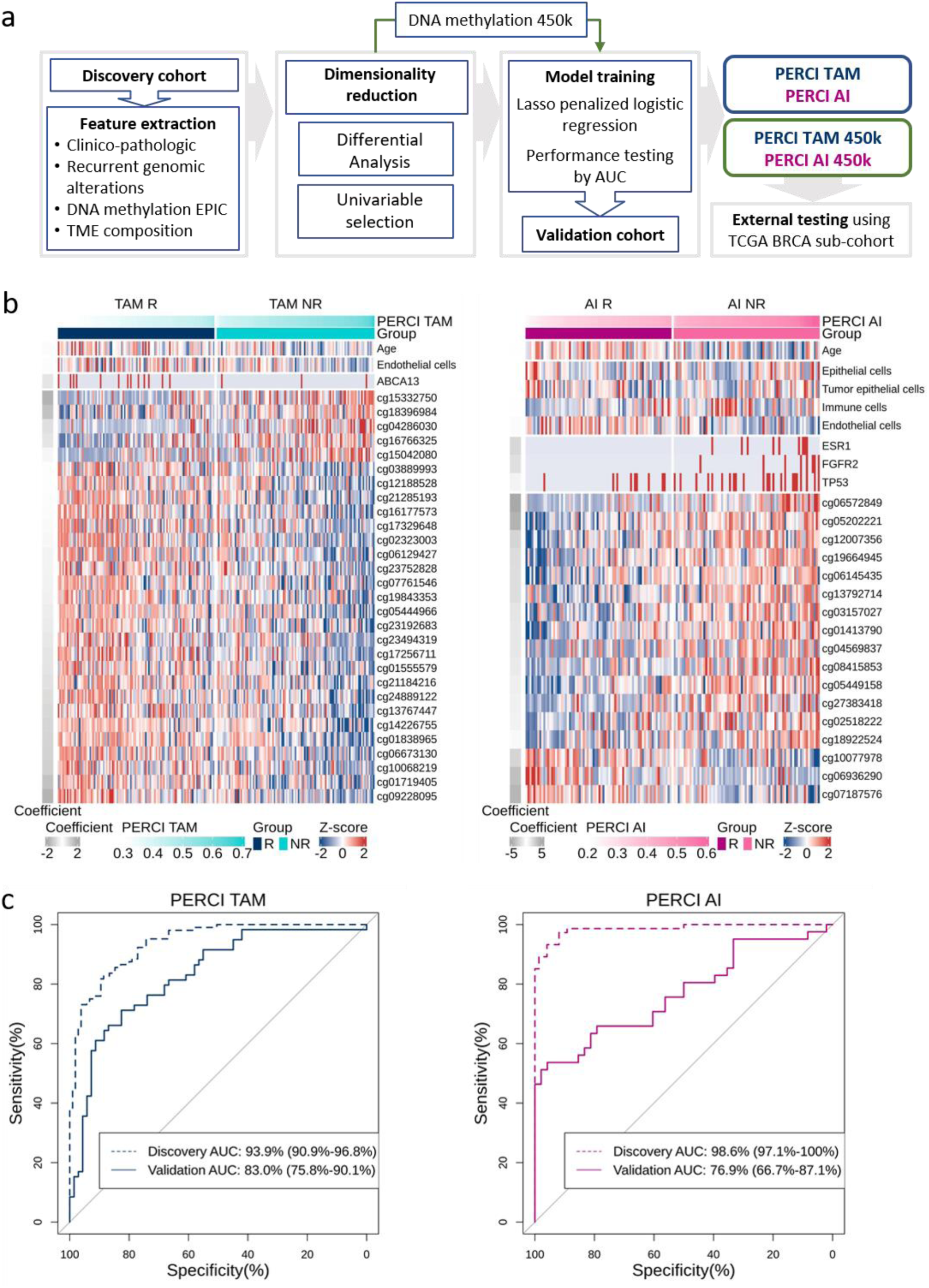
Developing the ‘Predictive Endocrine ResistanCe Index’ PERCI. (a) Workflow for the development of the Predictive Endocrine ResistanCe Index (PERCI). (b) Heatmap of z-scores for features selected to construct PERCI TAM (left panel) and PERCI AI (right panel). The coefficients on the right indicate the weights of each feature. (c) Area under the receiver operating characteristic curve (AUC) analysis of PERCI performance in the discovery and validation cohorts. The x-axis shows the specificity, while the y-axis shows the sensitivity. AUC with 95% confidence intervals.

**Table 1:**
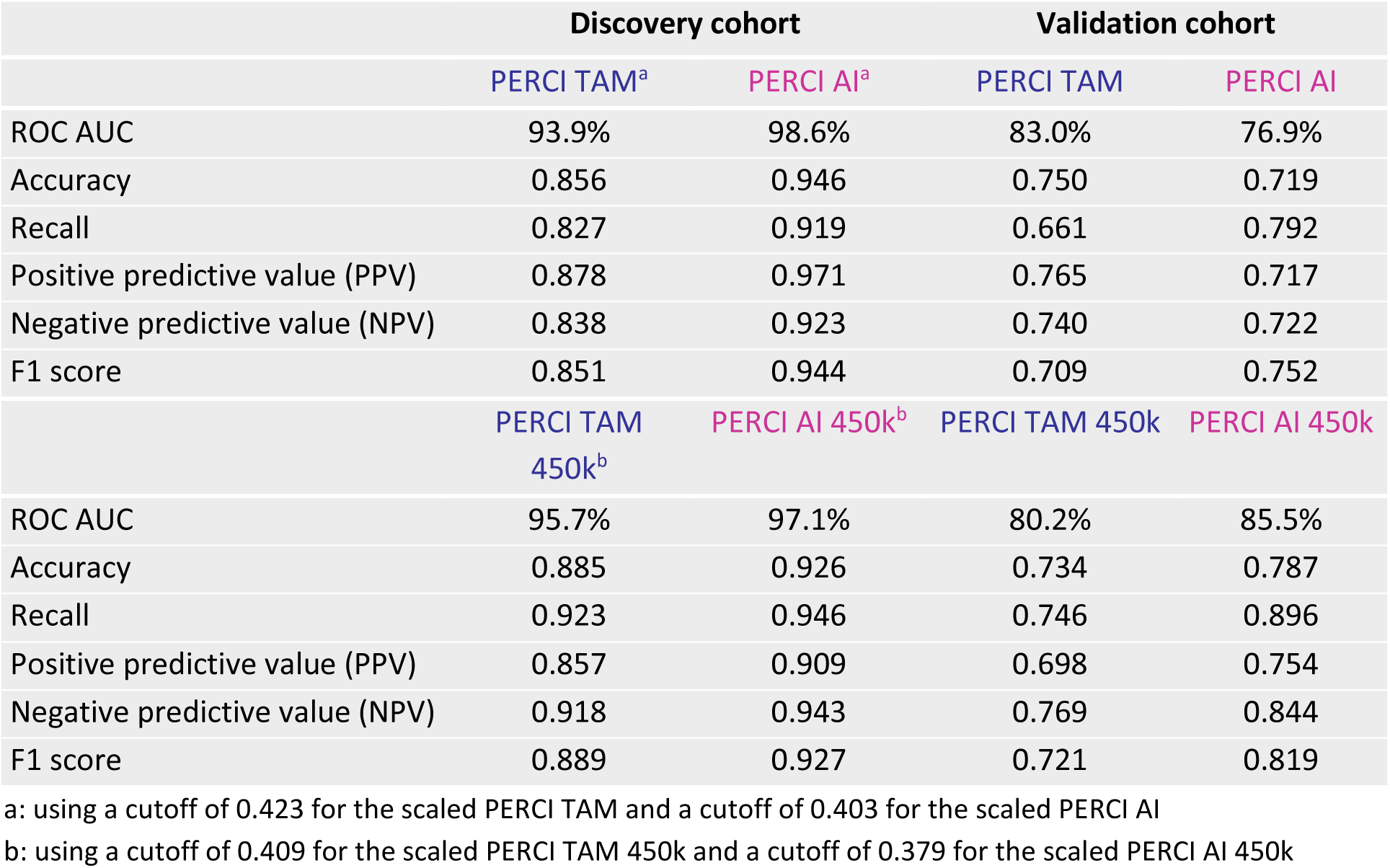
Performance of PERCI and PERCI 450k in the discovery and validation cohorts.

PERCI AI includes patient age, *ESR1*, *FGFR2*, and *TP53* genomic alterations, normal and tumor epithelial, endothelial and immune cell proportions, and methylation information for 17 AI DMS, most of which gained methylation in the AI non-responder group (**Fig. 5b, right**). Hypermethylated cg18922524, located in the promoter region of the homeobox transcription factor *HOXC4*, was exceptional as it was the CpG site with the strongest positive correlation with *HOXC4* mRNA levels (pearson correlation r = 0.475, fdr-corrected p-value *= 1.13E-15*) out of 12 AI DMS associated with the same gene (**Supp. Table S3, S5**), indicating increased gene expression. The individual CpG sites selected for PERCI AI reached ROC AUC values of 67.1 to 74.4% (**Supp. Fig. S5**), and all features combined stratified the NR and R groups with an AUC of 98.6% and excellent accuracy and true positive rate (PPV) (**Fig. 5c**, right panel; **Table 1**). As with PERCI TAM, the specificity of correctly annotating NR was higher than the sensitivity. In the validation cohort, we obtained an AUC of 76.9%. In this cohort, PERCI AI was better at correctly predicting the responder group.

Both PERCI TAM and PERCI AI correlated strongly with post-pET histology grade and Ki67 staining, with Spearman correlation coefficients ρ ranging from 0.55 to 0.78 in the discovery cohort (fdr-corrected p-values *< 0.01*) (**Supp. Fig. S2a**) and from 0.42 to 0.64 in the validation cohort (fdr-adjusted p-values *< 0.01*) (**Supp Fig. S2b**). Of note, in the discovery cohort, PERCI TAM and PERCI AI did not correlate with the Oncotype DX recurrence score (**Suppl. Fig. S2a**).

In summary, our novel predictors of resistance to TAM and AI therapy, PERCI TAM and PERCI AI, combine information on genomic alterations, patient age, TME composition and differential methylation, and with ROC AUCs of 83% for TAM and 76.9% for AI showed very good performance in the validation cohort.

### Adapting PERCI to the Illumina 450k platform and validation in the TCGA-BRCA cohort

The multi-criteria nature of PERCI may limit its applicability in clinical settings where not all data modalities may be readily available. For this reason, we trained a more streamlined version of PERCI, following the same approach as described above, but using only methylation data and age as input. In view of the widely available 450k Illumina array data, we restricted CpG sites to those that overlapped between EPIC and 450k arrays, with mean methylation differences between the R and NR groups in the ADAPT discovery cohorts of at least 5% and various significance thresholds (Methods). PERCI TAM 450k contains age and methylation information for 41 CpG sites, most of which were again hypomethylated in the TAM-NR group (**Supp. Fig. S6a**). Although the feature selection was independent, ten CpGs overlapped with PERCI TAM features. For PERCI AI 450k, in addition to age, 19 predominantly hypermethylated CpG sites were selected, eight of which overlapped with PERCI AI features (**Supp. Fig. S6b**). Both 450k-compatible versions of PERCI performed as well as or better than PERCI in the ADAPT cohorts, with ROC AUC >95% in the discovery and >80% in the validation cohorts (**Table 1, Supp. Fig. S6c**).

To test these classifiers in an external cohort, we used published TCGA-BRCA data and selected 296 cases with methylation data available and matching our cohort selection criteria (Methods). Patients were stratified by menopausal status in a TAM-like cohort (n=75) and an AI-like cohort (n=194) (clinico-pathologic characteristics in **Supp. Table S2, S6**). We divided each cohort into PERCI 450k high and low groups, using progression-free survival within 10 years as a surrogate for information on response to endocrine therapy. The group with high PERCI TAM 450k had 49 cases, whereas the low group had 26 cases (**Fig. 6a**, left panel). A Kaplan-Meier plot exhibited excellent stratification of these subgroups of treatment-naive cases. Patients in the PERCI TAM 450k low group had a favorable prognosis, as there were no instances of disease progression within the span of 10 years (log-rank p value = 0.03, **Fig. 6b**, upper panel). In addition, the PERCI TAM 450k low group had a significantly higher number of cases with the ILBC subtype compared to the PERCI TAM 450k high group (p=0.032, **Fig. 6c**, **Supp. Table S2, S6**), confirming our observations in the validation cohort (**Fig. 2h**).

**Fig. 6.**
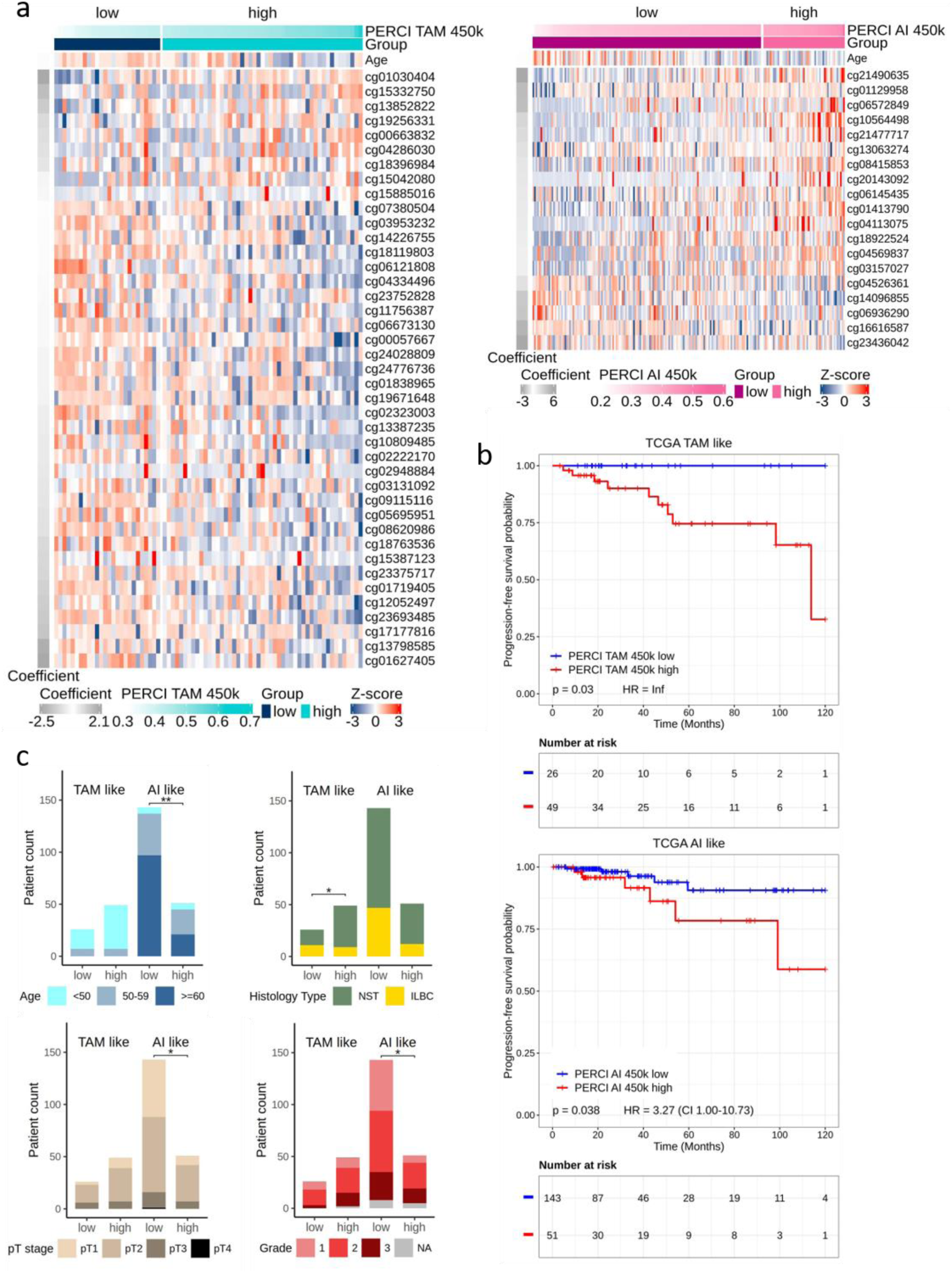
PERCI 450k predicts progression-free survival in the TCGA-BRCA sub-cohort. (a) Heatmap of selected features to construct PERCI TAM 450k (left panel) and PERCI AI 450k (right panel) in the TCGA-BRCA sub-cohort. (b) Kaplan-Meier curves of progression-free survival in the TCGA-BRCA sub-cohort on the basis of PERCI TAM 450k (upper) and PERCI AI 450k scores (lower). Cases were divided into high and low groups (blue: low, good prognosis, red: high, poor prognosis) using a cutoff of 0.424 for the scaled PERCI TAM 450k and a cutoff of 0.4011 for the scaled PERCI AI 450k. P values were calculated using the log-rank test. Hazard Ratios (HR) and their 95% CIs were estimated by an univariate proportional hazards regression. (c) Comparative clinical pathology of PERCI 450k low and high groups in the TCGA-BRCA sub-cohort. Statistical differences of variables were analyzed using Fisher’s exact test with *, ** p-value < 0.05, 0.01.

The PERCI AI 450k high group (**Fig. 6a**, right panel) consisted of 51 cases with a significantly worse prognosis than low group, with a hazard ratio of 3.27 for disease progression (log-rank *p=0.038*, **Fig. 6b**, lower panel). Cases in the PERCI AI 450k high group were younger (*p=1.9e-03*) and had higher stages (*p=0.026*) and histologic grades (*p=0.026*) compared to the PERCI AI 450k low group (**Fig. 6c, Supp. Table S2, S6**).

In conclusion, these data suggest that PERCI may not only be a predictor of primary endocrine resistance but may also have prognostic value.

## Discussion

The clinical definition of endocrine resistance implies progression or relapse of BC during endocrine therapy over a period of six months to two years [39]. If a test for primary endocrine resistance were available prior to initiation of endocrine therapy, inadequate therapy could be avoided or substituted. Consequently, this study aimed to characterize and compare molecular alterations associated with primary resistance to tamoxifen and aromatase inhibitor treatment and to use these features to develop classifiers for predicting treatment response and resistance.

Biologically, primary endocrine resistance can be assessed by a diminished or absent proliferative response of breast cancers to short-term preoperative endocrine therapy, as evidenced by *in situ* detection of the strictly proliferation-associated nuclear Ki67 antigen [9,12,40]. The considerable heterogeneity of luminal BC requires the analysis of a large number of cases. Our study included a total of 634 patients and had greater power to detect significant differences between response groups than earlier smaller studies using a similar approach [41,42]. Our study design was unique as it used tumor tissue from a unique prospective clinical trial (ADAPT) and different selection criteria for a discovery and a validation cohort. More specifically, the discovery cohort included equal numbers of responders and non-responders to pET, contrary to previous studies in this field [32,41]. In addition, responders and non-responders were precisely matched for several clinicopathologic characteristics to exclude confounding effects related to differences in baseline tumor characteristics between responders and non-responders. For example, both responders and non-responders had equal proportions of G3-differentiated BCs (**Fig. 2a, Supp. Table S1a**). Thus, subsequent molecular analyses were informative of pET response determinants and were not biased by dominant molecular features associated with G3 differentiation. In addition, the criteria for endocrine response and non-response were particularly stringent (see selection criteria). In contrast, the validation cohort had more relaxed selection criteria and was more representative of the patient populations encountered in clinical practice (**Fig. 2g-l, Supp. Table S1b**). Therefore, in combination, our study design was well suited for the identification of informative markers of pre-operative treatment failure in the discovery cohort and their validation under representative clinical conditions in the validation cohort.

The mechanisms driving the development of primary endocrine therapy resistance can be diverse [43]. To develop our Predictive Endocrine Resistance Indices (PERCI), we combined clinico-pathologic data with data on significant recurrent genomic alterations and epigenetic changes associated with pET resistance after three weeks of TAM or AI treatment. To select the most informative features, we used a machine learning approach based on lasso penalized logistic regression [44] (**Fig. 5**). For PERCI AI, the algorithm selected genomic alterations in *ERS1, FGFR2* and *TP53*, key cancer driver genes that were enriched in AI non-responders, while PERCI TAM was based on a higher mutation frequency of *ABCA13* in TAM responders.

Somatic mutations linked with primary resistance are commonly detected through their prevalence in metastatic lesions when compared to primary breast cancer [45]. Considerable progress was achieved by the discovery of *ESR1* mutations, which are only rarely found in primary luminal cancers (<1%) but are enriched in up to 15%-30% of cases in metastatic luminal cancers during adjuvant anti-endocrine therapy and lead to ligand-independent autocrine growth of tumor cells. In our discovery cohort, we detected *ESR1* mutations and amplifications in 12% of AI non-responders, but in none of AI responders. Consistently, Ferrando *et al.* detected *ESR1* amplifications enriched in metastatic lesions of BC cases as compared to primary tumors exclusively in patients treated with adjuvant AI, but not in TAM-treated patients [46]. *FGFR2* amplifications were also enriched predominantly in AI non-responders. Mao et al. previously reported an increase in *FGFR2* amplification in post-resistance biopsies treated with ER-targeted therapy [47]. Enrichment in metastatic luminal BC has also been demonstrated for *TP53* mutations, with an incidence of more than 25% in metastases [45]. As reported recently by us [15], our method to identify primary resistance by evaluating the lack of proliferative response of breast cancer to short-term endocrine therapy revealed *TP53* to be the most commonly mutated gene associated with therapy failure in the discovery cohort (avg. mutation rate of 15%)(**Fig. 3c**). These findings were confirmed for both treatment regimens in the validation cohort (**Fig. 3d**). These results suggest that the two approaches to assessing mechanisms of endocrine resistance yield overlapping information and support the relevance of these RGA as features in PERCI. In line with this, Gellert et al. reported reduced suppression of Ki67 within the poor responder group for *TP53* mutant tumors in ER-positive BC treated with AI for 2 weeks [41]. As *TP53* mutations lead to aberrant nuclear accumulation of the mutant p53 protein [48], p53 IHC may potentially be used as a surrogate marker for endocrine resistance in a clinical setting [36].

In contrast to the genomic alterations included in PERCI AI, mutations in *ABCA13* (ATP Binding Cassette Subfamily A Member 13), which were included as a feature in PERCI TAM, indicated better response to treatment (**Fig. 3c**). ABC transporters, such as ABCA13, contribute to the development of therapy resistance by ATP-dependent drug efflux [49]. Therefore, inactivation through point mutations could potentially enhance therapy response. Gellert et al. reported a slightly elevated (but statistically insignificant) prevalence of *ABCA13* mutations in patients responsive to AI treatment [41].

Another interesting gene with mutations enriched in the TAM responder group, which was not included in PERCI TAM, is *CBFB* (Core-Binding Factor Subunit Beta) (**Fig. 3c,d**). The *CBFB* gene encodes the RUNX transcriptional coregulator CBFβ, which has an important function in transcriptional and translational regulation [32,50]. With a frequency of 17.9% and 22.2% of CBFB mutations in the discovery and validation cohorts, our results showed a significant increase in splicing and frameshift mutations compared to TAM non-responders (**Fig. 3c,d**).

Consistently, a recent analysis of *CBFB* mutation patterns in two large breast cancer cohorts (METABRIC, TCGA-BRCA) concluded that *CBFB* mutations are associated with improved survival in HR+/HER2-patients [51], although the underlying mechanisms are not completely understood [52].

In addition to genomic alterations and age, PERCI TAM and PERCI AI included DNA methylation differences between responders and non-responders and epigenetically informed cell type composition (**Fig. 4,5**). CpG sites selected for PERCI TAM were mainly lower methylated in TAM non-responders. Global methylation loss in large regions of heterochromatin represents one of the hallmarks of cancer epigenomes and is mediated by increased cell proliferation [27,37]. In contrast, cg16766325 in the *SPRY2* promoter region was hypermethylated in TAM non-responders, indicative of SPRY2 downregulation (**Supp. Table S5**). SPRY2 inhibits cell proliferation by acting as a feedback inhibitor of the RAS-MAPK pathway downstream of FGF/FGFR [53], and loss of *SPRY2* expression was recently shown to promote cancer-associated fibroblast activation and BC progression [54]. In line with these findings, KRAS signaling, apical cell-cell junctions, and epithelial-mesenchymal transition (EMT) were identified as the most enriched gene sets associated with differential methylation in the TAM group (**Supp. Fig. S3**) and might promote TAM resistance [43,55,56].

The DNA methylome associated with AI resistance was characterized by a predominant gain in methylation, in contrast to the prominent loss of methylation observed in TAM non-responders. Accordingly, AI DMS included as features in PERCI AI were mostly hypermethylated in AI non-responders. Interestingly, for a subset of these hypermethylated genes methylation positively correlated with gene expression. Many of these hypermethylated, upregulated genes belonged to the family of developmental transcription factors [57] and have been previously implicated in breast cancer etiology and the development of endocrine resistance [58], including *GATA2* [59], *HOXC4* [60], *HOXB13* [61], *HOXC13* [62], *MNX1* [63], *OTX1* [64], *PAX7*, *SOX2* [65] and *WT1* [66]. Notably, one of the 12 AI DMSs associated with *HOXC4* was included in both AI models. Consistent with our findings, *HOXC4* hypermethylation was identified as a biomarker of endocrine resistance in a study analyzing a small subset of 31 TCGA-BRCA cases that had received endocrine therapy [67]. In addition, the authors identified epithelial-stromal interaction 1 (*EPSTI1*) promoter hypermethylation associated with endocrine resistance, and two AI DMSs (cg01536987, cg22905097) overlapped with the differentially methylated region identified in this previous study. EPSTI1 is overexpressed in aggressive breast cancer and may confer breast stem/progenitor cell properties [68]. In our study, we detected weak inverse correlation of cg01536987 methylation with gene expression in the TCGA-BRCA sub-cohort (**Supp. Table S5**).

Gene set overrepresentation analyses indicated that AI-DMS-associated genes were enriched in gene sets related to hypoxia [69] and estrogen response (**Supp. Fig. S3d**). These findings suggest that resistance to AI therapy may be due to an altered response to hypoxic conditions. In a previous report by Oshi *et al*., three months of neoadjuvant AI was shown to reduce expression of hypoxia-inducible factor-1 (*HIF-1*), a master regulator of oxygen homeostasis [70]. Oshi *et al.* also linked low expression of early estrogen response genes to a reduced response to endocrine therapy [70]. Consistent with these findings, Jeong et al. demonstrated that re-expression of the epigenetically silenced early estrogen response gene *ELOVL2* rescued its downstream signaling and TAM sensitivity in TAM-resistant MCF7 cells and in a xenograft mouse model [71]. In our study, cg14153064 located in the promoter region of *ELOV2* was significantly hypermethylated in AI non-responders (**Supp. Table S5**).

Given that DNA methylation is a stable epigenetic mark of cellular identity [72], we used reference DNA methylomes to infer bulk tumor and tumor microenvironment composition. As experimental validation, we observed high correlation of bioinformatically-estimated tumor-infiltrating lymphocytes (TIL) proportions with pathologically-determined TIL levels in both cohorts (**Supp. Fig. S3**). Proportions of normal and tumor epithelial cells, immune cells, and endothelial cells, were selected as features of PERCI AI. The proportions of normal and tumor epithelial and immune cells did not differ between responders and non-responders and had only minor contributions to the model. However, because bulk methylation values are affected by tumor purity and tumor purity thus indirectly affected PERCI scores, samples with high normal and tumor epithelial cell content (and conversely low immune cell content) had the lowest and highest PERCI AI scores in the responder and non-responder groups, respectively. We also noticed that samples with *TP53* mutations seemed to be enriched in immune cells, indicating that there might be some interaction between features.

A limitation to the use of our classifiers in clinical settings may be sample size, quality of DNA from FFPE tissue, or instrumentation and budget constraints that limit the generation of high-quality NGS data to identify mutation status for the genes included in the classifiers [73]. To circumvent this potential limitation, we generated a simplified version of PERCI, PERCI 450k, based only on DNA methylation and age (**Supp. Figure S6**). The feasibility of using DNA methylation as a cost-effective classifier in clinical samples has been demonstrated in brain tumors [74], where methylation-based profiling has now been incorporated into the WHO classification of central nervous system tumors [75]. In our cohorts, PERCI 450k performed as well as or better than PERCI in stratifying responders and non-responders, with ROC AUCs above 80% even in the validation cohort (**Table 1**). In addition, PERCI 450k showed promise as a prognostic marker in predicting progression-free survival in the TCGA-BRCA sub-cohort (**Fig. 6**).

## Conclusions

This study has shown that the cellular pathways of endocrine resistance to TAM and AI are affected by distinct genetic and epigenetic alterations. The delineation of differences between the mechanisms of endocrine resistance between the two mainstays of standard endocrine therapy in BC opens the perspective of overcoming it by switching from one drug to the other. PERCI as a biomarker of endocrine resistance can be readily used for risk stratification in future therapeutic trials in BC.

## Declarations

### Ethics approval and consent to participate

The study design is following the guidelines of the local ethics committee (Ethics committee of the Medical School Hannover, ID 2716–2015).

## Consent for publication

Not applicable.

## Availability of data and materials

The datasets used and analyzed during the current study are available from the corresponding author on reasonable request.

## Authors’ contributions

H.K., N.H., U.M. and M.C. designed the study. O.G., M.G., S.K., U.N., and N.H. collected patient samples and data. R.K. and M.C. selected the patients and performed the matching. M.C., H.C., L.D.K, M.R. performed the immunohistochemical stainings and pathology assessment. S.B., G.Z., C.G., V.J., D.S., U.L., W.H., B.S., U.M. and J.B. performed the experiments, analyzed, and interpreted the data. U.M. provided supervision. G.Z. and C.G. wrote the manuscript. S.B., M.C. H.K., C.P., N.H. and U.M. reviewed and edited the manuscript. All authors read and approved the final manuscript.

## Data Availability

All data produced in the present study are available upon reasonable request to the authors.

## List of abbreviations

ADAPT: adjuvant dynamic marker-adjusted personalized therapy
AI: aromatase inhibitors
AUC: area under curve
BC: breast cancer
DMS: differentially methylated CpG site
EMT: epithelial –mesenchymal transition
EPIC: Infinium Methylation
EPIC: bead chip
ER: estrogen receptor
FFPE: Formalin-fixed paraffin-embedded
GSEA: gene set enrichment analysis
H&E: haematoxylin and eosin
ILBC: intralobular breast cancer
Ki67: proliferation marker Ki67
MAF: minor allele frequency
MeTILs: DNA methylation-based proportions of tumor infiltrating lymphocytes
MHH: Medizinische Hochschule Hannover
MSigDB: Molecular Signatures Database
NGS: next-generation sequencing
NPV: negative predictive value
NR: non-responder
PaTILs: pathological-assessed tumor infiltrating lymphocytes
PERCI: Predictive Endocrine ResistanCe Index
pET: preoperative endocrine therapy
PFS: progression-free survival
pN: pathologic lymph node status
PPV: positive predictive value
PR: progesterone receptor
pT: pathologic tumor stage
R: responder
RGA: recurrent genomic alteration
ROC: Receiver Operating Characteristics
RS: Oncotype DX Recurrence Score
SNP: single nucleotide polymorphism
TAM: tamoxifen
TCGA-BRCA: The Cancer Genome Atlas Breast Cancer study
TILs: tumor-infiltrating lymphocytes
TME: tumor microenvironment
TP53: tumor suppressor protein p53
TSS: transcription start site
WSG: West German Study Group

## Acknowledgements

We thank the patients and families who contributed to this study. The authors acknowledge the DKFZ Genomics and Proteomics Core Facilities for excellent service. We would like to thank Annette Weninger and Karin Klimo (German Cancer Research Center) for excellent technical assistance. The results reported here are in part based upon data generated by *The Cancer Genome Atlas* managed by the NCI and NHGRI. Information about TCGA can be found at http://cancergenome.nih.gov.

## Authors’ information (optional)

### Additional files

**Supp. Fig. S1 (pdf). Endocrine therapy-induced changes in histologic grade** per treatment group in the discovery and validation cohorts.

**Supp. Fig. S2 (pdf): Correlations of clinical features with TME components, PERCI scores and mutations**. Spearman and Kendall correlation coefficients were calculated between selected features in the discovery and validation cohort for the TAM and AI groups.

**Supp. Fig. S3 (pdf): pET resistance-related alterations in the methylome and tumor microenvironment**. Density plots of mean methylation beta values of responder vs. non-responder groups for TAM– and AI-treated cases in the discovery and the validation cohort. Percentage of DMS, split in hypo-and hypermethylated in NR, overlapping with chromatin regions derived from ChromHMM analyses of MCF7 cells. Spearman correlation of stromal TILs from H&E-stained slides with methylation-derived TILs. Distributions of major immune cell fractions in responder vs. non-responder groups for TAM– and AI-treated cases in the discovery and the validation cohort.

**Supp. Fig. S4 (pdf): ROC AUC of all predictors of PERCI TAM**. Area under the receiver operating characteristic curve (AUC) of the individual predictors of PERCI TAM in the TAM discovery cohort.

**Supp. Fig. S5 (pdf): ROC AUC of all predictors of PERCI AI**. Area under the receiver operating characteristic curve (AUC) of the individual predictors of PERCI AI in the AI discovery cohort.

**Supp. Fig. S6 (pdf): Performance of PERCI TAM 450k and PERCI AI 450k in the discovery and validation cohorts**. Heatmap of z-scores of age and methylation of CpG sites selected to build PERCI TAM 450k and PERCI AI 450k. Area under receiver operating characteristic curve (AUC) analysis of PERCI 450k’s performance in the discovery and validation cohorts.

**Supplementary Table S1 (docx). Clinico-pathologic characteristics of the discovery cohort (a) and the validation cohort (b).**

**Supplementary Table S2 (docx). Clinico-pathologic features of the TCGA-BRCA sub-cohort.**

**Supplementary Table S3 (xlsx): Clinico-pathologic data of the discovery and validation cohorts and detailed description of the Predictive Endocrine ResistanCe Index (PERCI) and PERCI 450k.** Clinicopathological information, cell type composition, data availability indicator in the discovery and the validation cohort. Features, coefficients and data of PERCI TAMl, PERCI TAM 450k, PERCI AI and PERCI AI 450k, PERCI scores.

**Supplementary Table S4 (xlsx): Detailed information of the panel sequencing**. Sequencing information, and details of mutations and copy number status for cases of the discovery and the validation cohorts.

**Supplementary Table S5 (xlsx): Detailed information of the differentially methylated CpG sites (DMS)**. IDs, genomic positions, means per TAM and AI groups, statistic, annotation to genes and chromatin regions (MCF7), expression of annotated genes in TCGA-BRCA sub-cohort, pearson and spearman correlation coefficients and p-values, beta values for TAM DMS and AI DMS in the discovery and the validation cohorts and in the TCGA-BRCA sub-cohort (overlap with 450k), RNA expression values (log2 FPKM-UQ) of the annotated genes in the TCGA-BRCA sub-cohort.

**Supplementary Table S6 (xlsx): Clinico-pathological data of the TCGA-BRCA sub-cohort and PERCI 450k prediction models**. Clinicopathologic information. Features, coefficients and data of PERCI TAM 450k and PERCI AI 450k model in the TCGA-BRCA sub-cohort and PERCI 450k scores.

**Supplementary Fig. S1.**
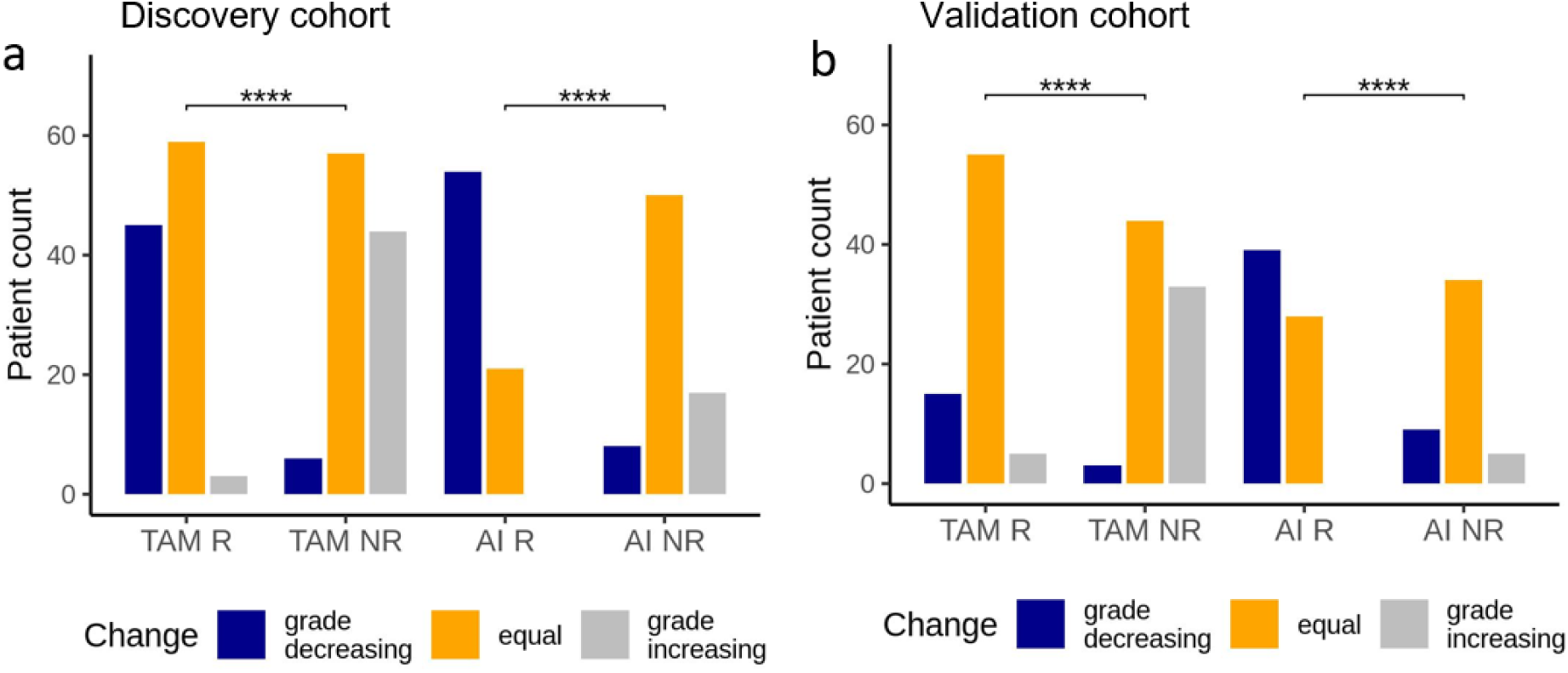
Endocrine therapy-induced changes in histologic grade from baseline to post-pET in the discovery (a) and validation cohort (b). Statistical differences tested using Chi-squared test with p-value **** < 0.0001.

**Supplementary Fig. S2.**
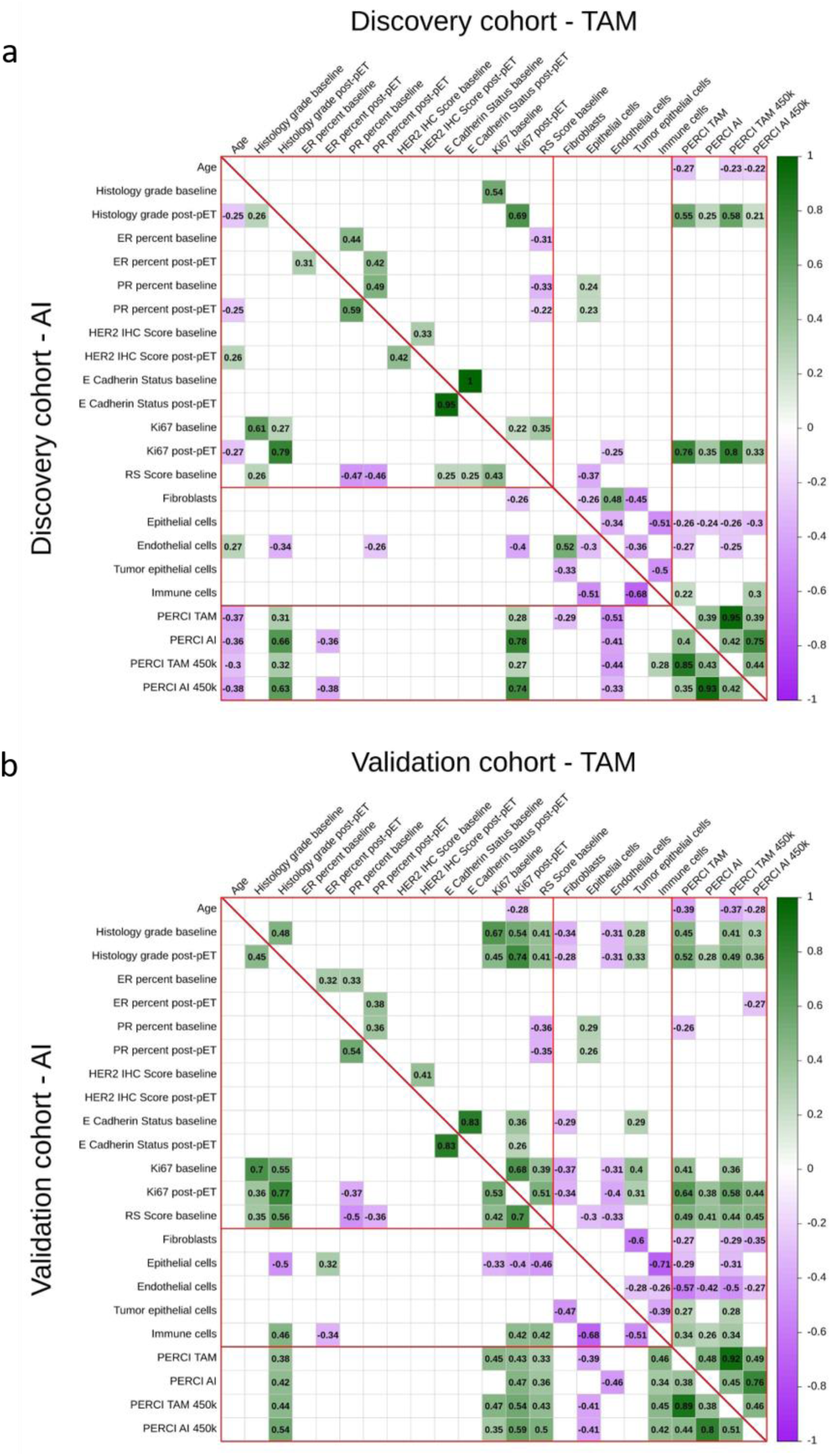
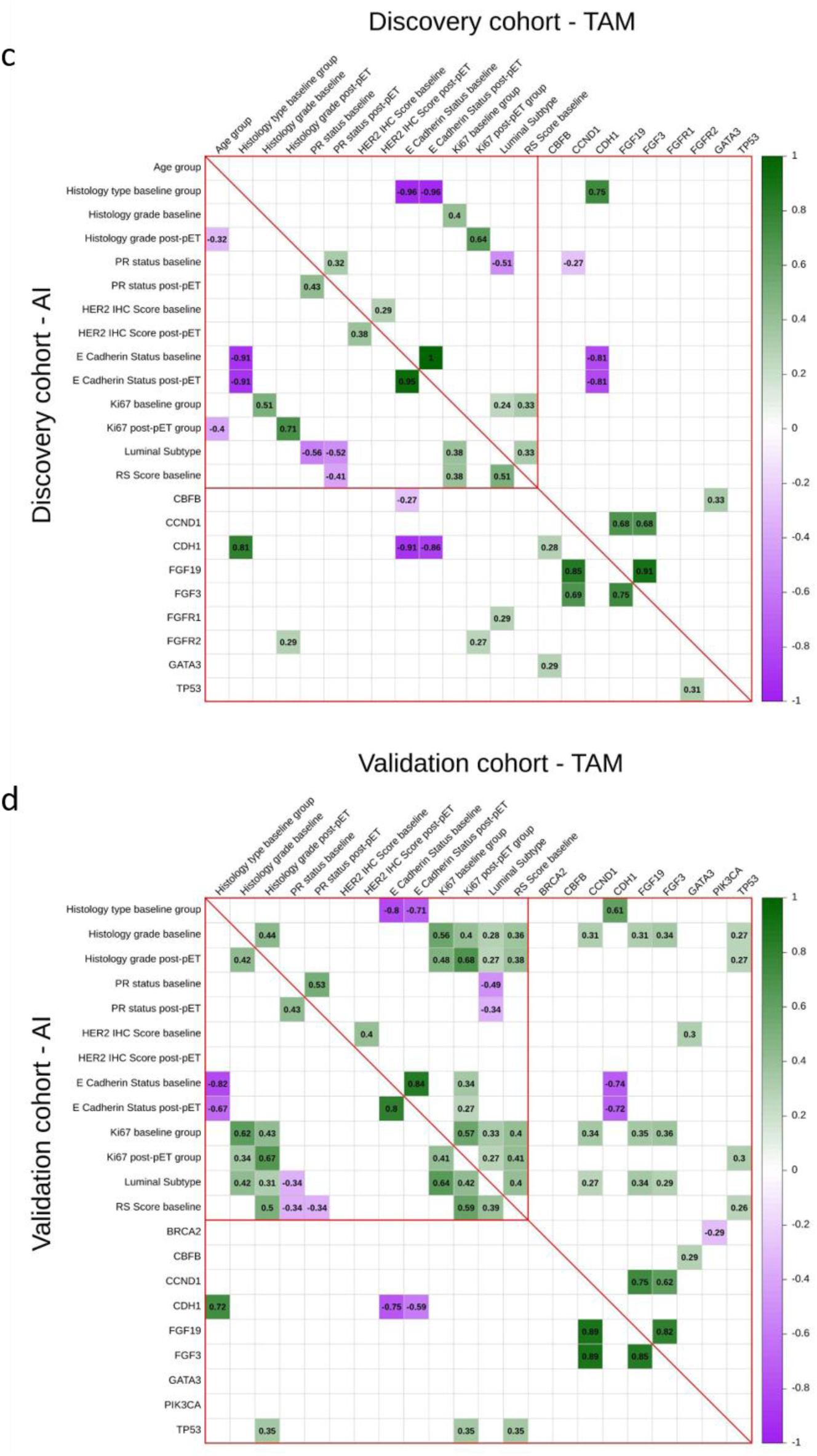
Correlations of clinical features with TME components, PERCI scores and mutations. Spearman correlation coefficients were calculated between selected features in the discovery (a) and validation cohort (b) for the TAM and AI groups. Correlations are indicated by a color gradient from purple (rho = –1) to green (rho = 1). (c,d) Kendall correlation for ordinal clinical data and mutation counts in the (c) discovery cohort, (d) validation cohort for the TAM and AI groups. Only results with statistically significant differences between response groups are shown (two-tailed test of significance which compares the observed value of correlation coefficient to its expected value under the null hypothesis (no correlation between the two variables), fdr-corrected p-value *< 0.01*).

**Supp. Fig. S3:**
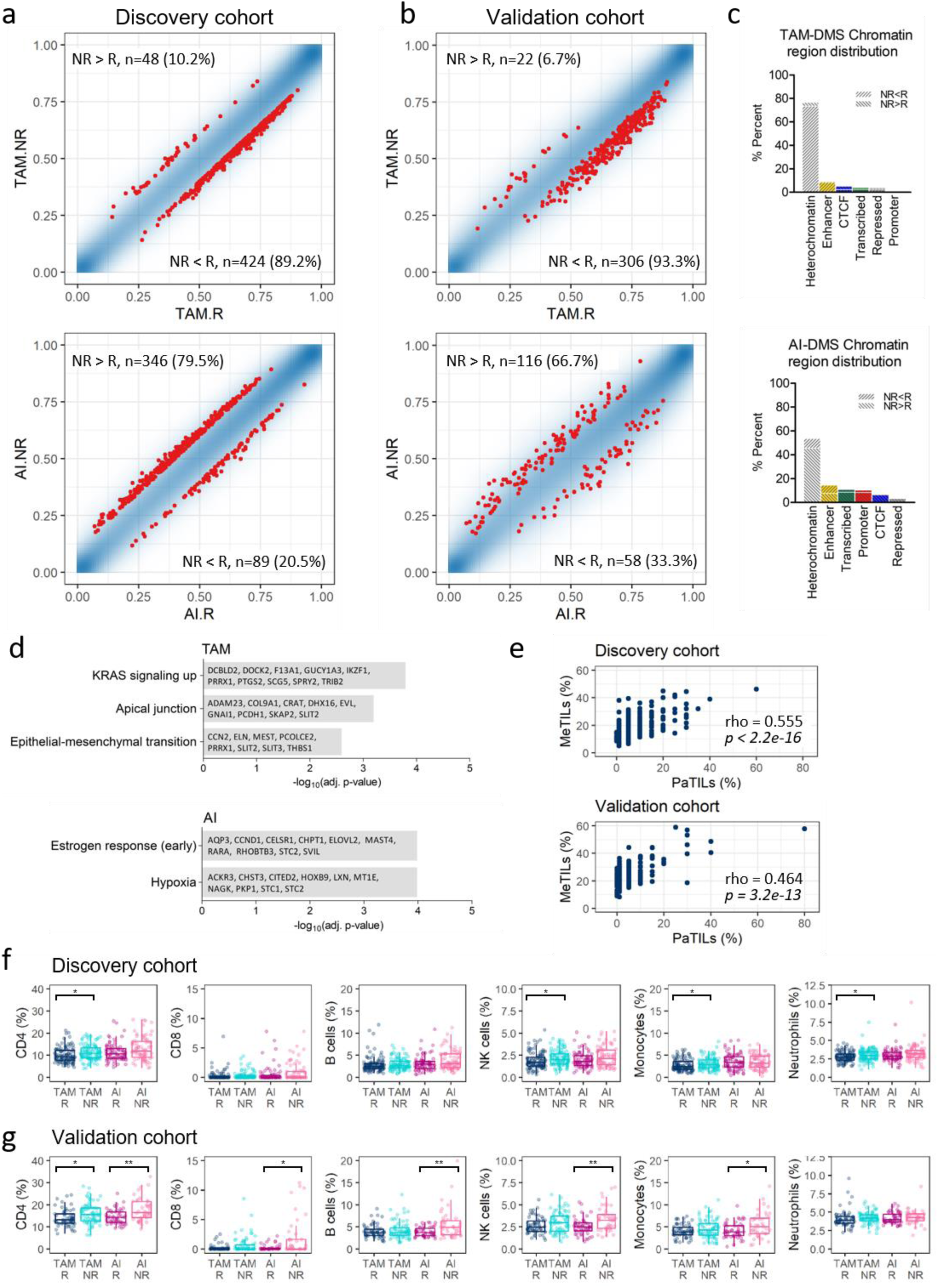
pET resistance-related alterations in the methylome and tumor microenvironment. (a) Density plots of mean methylation beta values of responder vs. non-responder groups (blue dots) for TAM-(upper panel) and AI-treated cases (lower panel) in the discovery cohort. Differentially methylated CpG sites (DMS) with a 10% methylation difference between R and NR (p < 0.005, limma) are highlighted in red (TAM: n=472 DMS, AI: n=435 DMS). Numbers of DMS with loss (NR < R) and gain in methylation in the NR group (NR > R) are indicated in the corners. (b) Density plots of mean methylation beta values of R vs. NR groups (blue dots) for TAM-(upper panel) and AI-treated cases (lower panel) in the validation cohort. DMS from (a) with a 5% methylation difference between R and NR (p < 0.05, limma) are highlighted in red (TAM: n=328 DMS, AI: n=174 DMS). (c) Percentage of DMS, split in hypo-and hypermethylated in NR, overlapping with chromatin regions derived from ChromHMM analyses of MCF7 cells (upper: TAM, lower: AI). (d) GSEA overrepresentation analysis of genes associated with TAM-DMS (upper) and AI-DMS (lower) in MSigDB hallmark gene sets. (e) Scatter plot depicting percentage of stromal TILs quantified by visual assessment on routine hematoxylin and eosin (H&E)-stained slides (PaTILs) vs. percentage of methylation-derived TILs (MeTILs, sum of CD4+, CD8+, B cell and NK cell percentages). Spearman’s rank correlation coefficient rho and p-value for the correlation between both methods is indicated. (f,g) Boxplots of major immune cell fractions calculated from methylation data using the reference-based Houseman algorithm (Methods) in the discovery (f) and validation cohort (g). R and NR groups per pET were compared using Wilcoxon test with *, **, *** fdr-adjusted p-value < *0.05, 0.01, 0.001*.

**Supp. Fig. S4:**
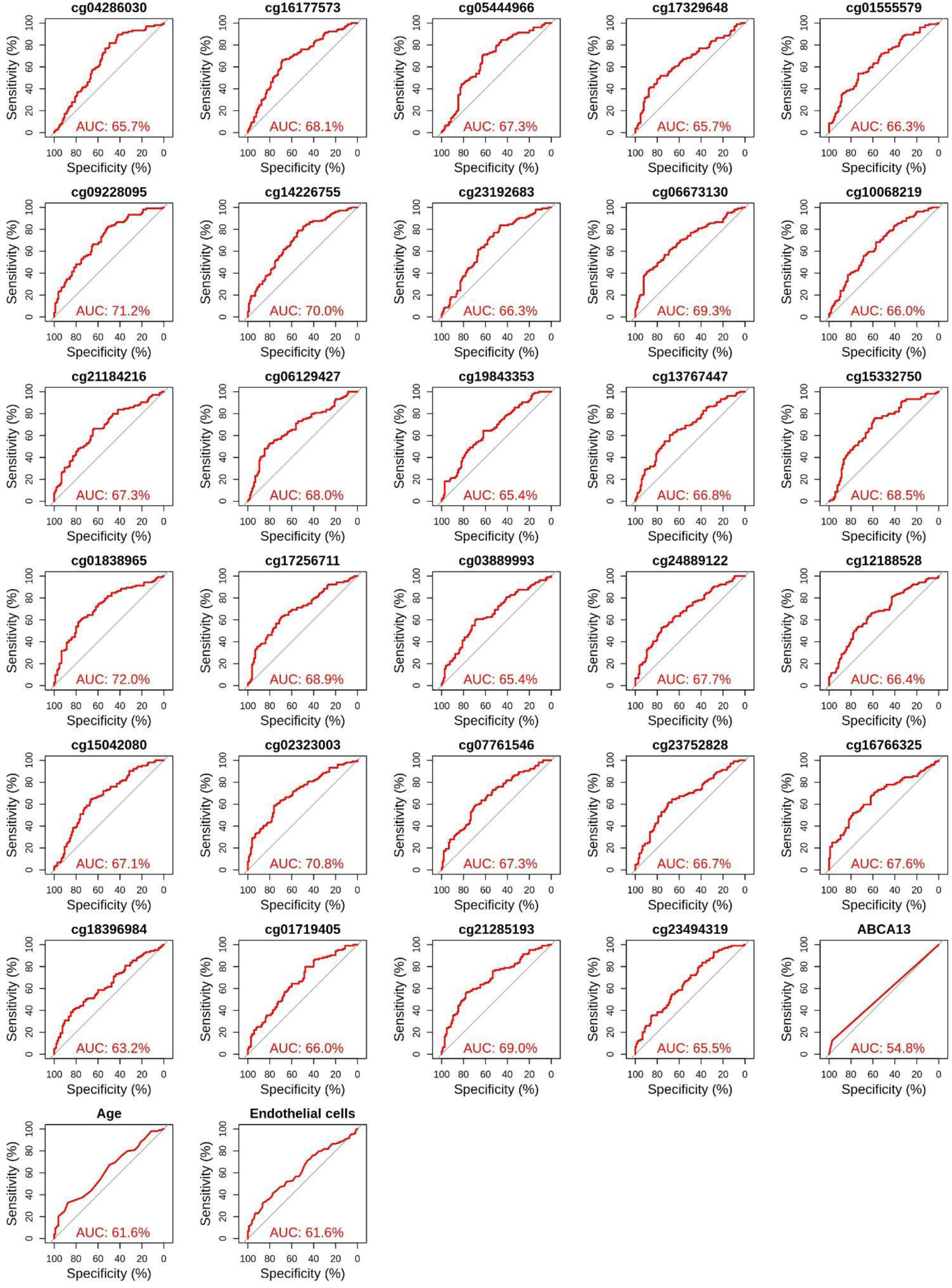
ROC AUC of all predictors of PERCI TAM. Area under the receiver operating characteristic curve (AUC) of the full model (upper left) and the individual predictors of PERCI TAM in the TAM discovery cohort. The x-axis shows the specificity, while the y-axis shows the sensitivity.

**Supp. Fig. S5:**
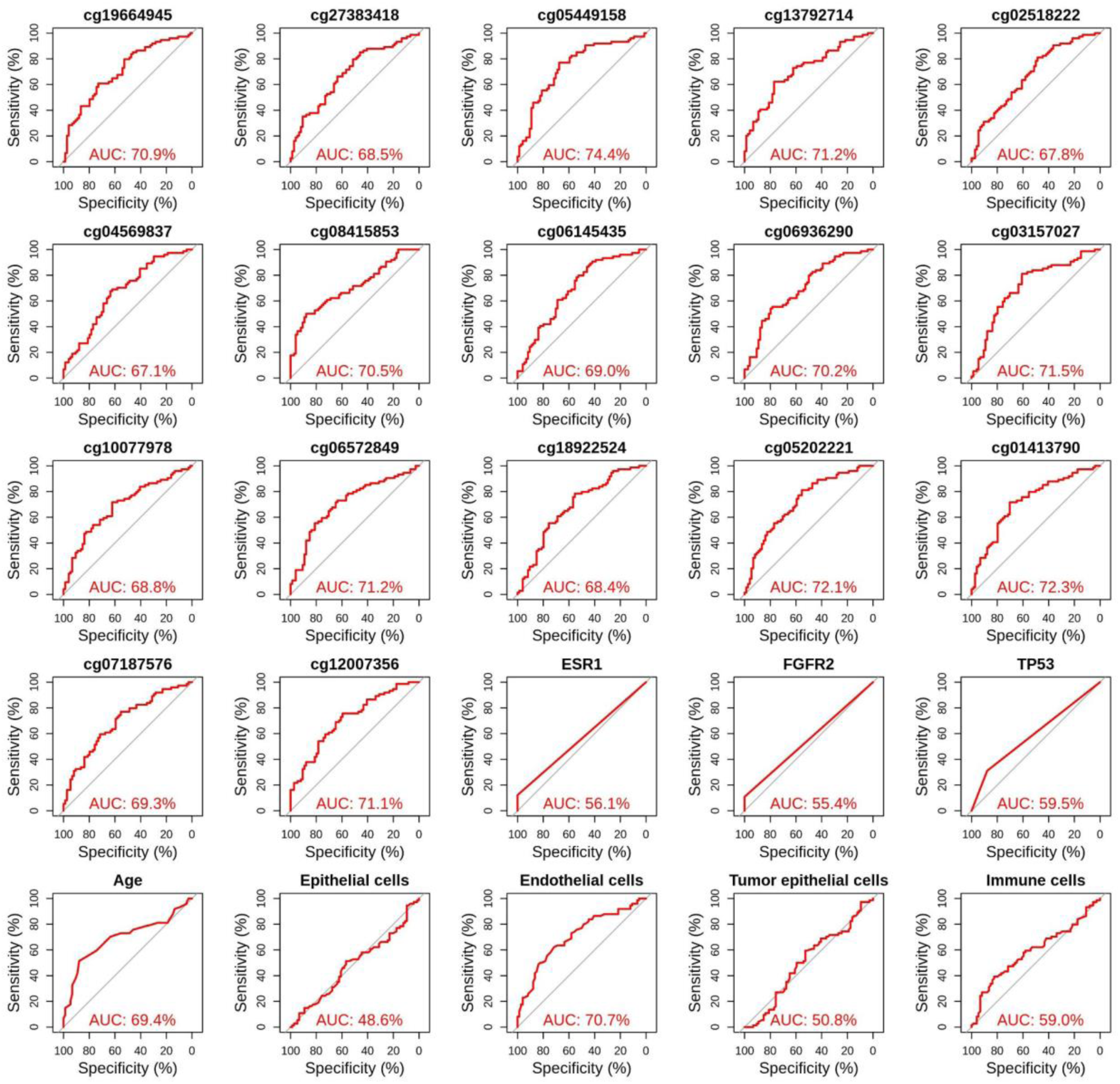
ROC AUC of all predictors of PERCI AI. Area under the receiver operating characteristic curve (AUC) of the full model (upper left) and the individual predictors of PERCI AI in the AI discovery cohort. The x-axis shows the specificity, while the y-axis shows the sensitivity.

**Supp. Fig. S6:**
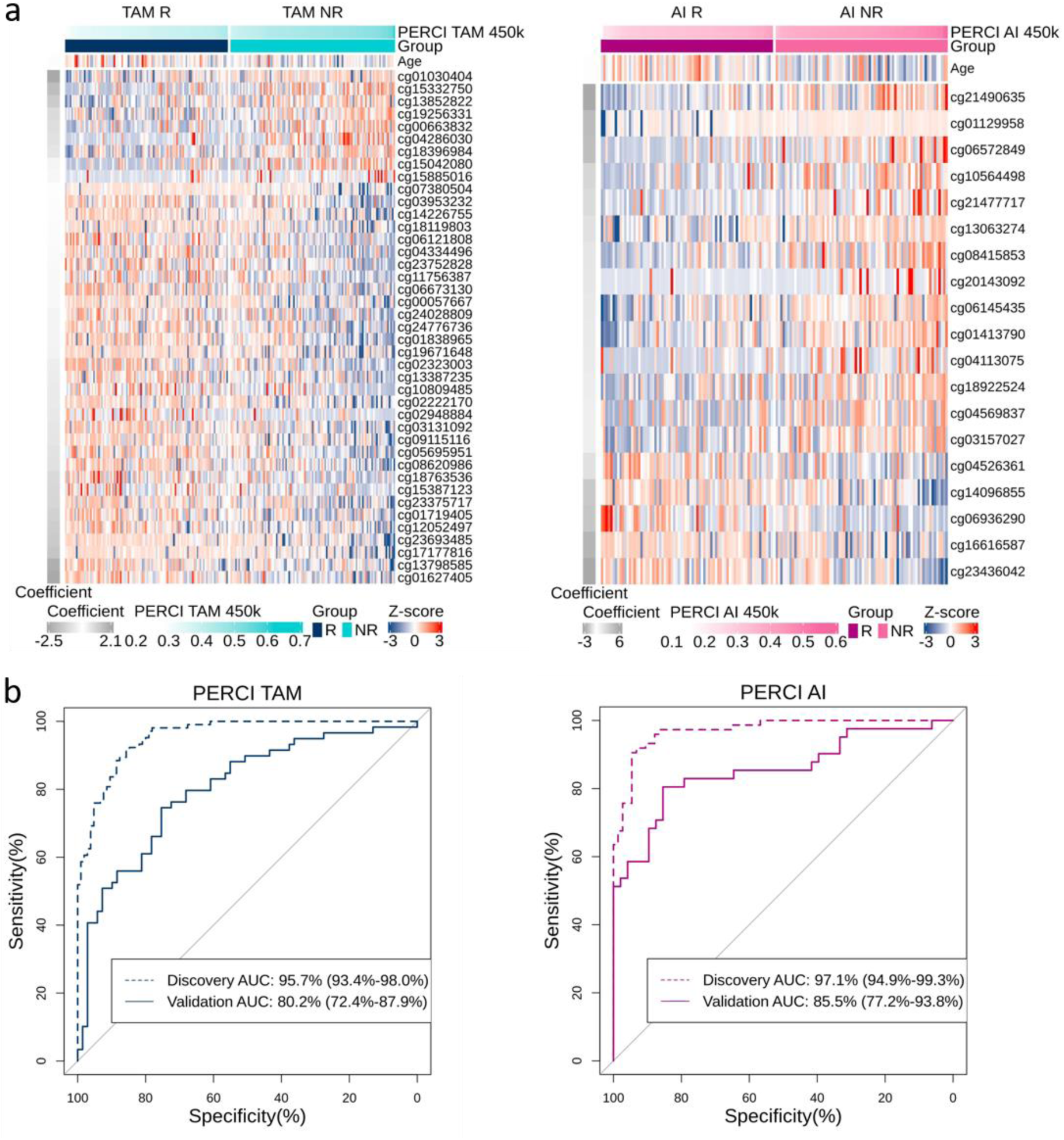
Performance of PERCI TAM 450k and PERCI AI 450k in the discovery and validation cohorts. (a) Heatmap of methylation z-scores of age and selected CpG sites to build PERCI TAM 450k (left panel) and PERCI AI 450k (right panel). (b) Analysis of the performance of PERCI 450ks in the discovery and validation cohorts by area under the receiver operating characteristic curve (AUC). The x-axis shows specificity and the y-axis shows sensitivity. AUC with 95% confidence intervals.

**Supplementary Table S1a.** Clinico-pathologic characteristics of the discovery cohort.

|  | All | TAM |  | AI |  |  |  |
| --- | --- | --- | --- | --- | --- | --- | --- |
|  |  | Responder<sup>c</sup> | Non-Responder<sup>b</sup> | Responder<sup>a</sup> | Non-Responder<sup>b</sup> |  |  |
|  | n=364 | n=107 | n=107 | n=75 | n=75 | *P*<sub>TAM</sub> | *P*<sub>AI</sub> |
|  | n (%) | n (%) | n (%) | n (%) | n (%) |  |  |
| age |  |  |  |  |  | **0.023** | **1.6e-05** |
| < 50 | 116 (32%) | 49 (46%) | 62 (58) | 0 | 5 (7%) |  |  |
| 50 - 59 | 133 (36%) | 47 (44%) | 43 (40%) | 9 (12%) | 34 (45%) |  |  |
| ≥ 60 | 115 (32%) | 11 (10%) | 2 (2%) | 66 (88%) | 36 (48%) |  |  |
| histology, baseline |  |  |  |  |  | **1** | **1** |
| NST | 324 (89%) | 92 (86%) | 92 (86%) | 70 (93%) | 70 (93%) |  |  |
| ILBC | 40 (11%) | 15 (14%) | 15 (14%) | 5 (7%) | 5 (7%) |  |  |
| pT stage |  |  |  |  |  | **0.566** | **1** |
| pT1 | 244 (67%) | 71 (66%) | 71 (66%) | 51 (68%) | 51 (68%) |  |  |
| pT2 | 118 (32%) | 35 (33%) | 35 (33%) | 24 (32%) | 24 (32%) |  |  |
| pT3 | 2 (1%) | 1 (1%) | 1 (1%) | 0 | 0 |  |  |
| pT4 | 0 | 0 | 0 | 0 | 0 |  |  |
| pN stage |  |  |  |  |  | **1** | **1** |
| pN0 | 342 (94%) | 102 (95%) | 102 (95%) | 69 (92%) | 69 (92%) |  |  |
| pN1+ | 22 (6%) | 5 (5%) | 5 (5%) | 6 (8%) | 6 (8%) |  |  |
| grade, baseline |  |  |  |  |  | **0.137** | **NA** |
| G1 | 20 (6%) | 7 (7%) | 7 (7%) | 2 (3%) | 2 (3%) |  |  |
| G2 | 198 (54%) | 73 (68%) | 73 (68%) | 27 (36%) | 27 (36%) |  |  |
| G3 | 146 (40%) | 27 (25%) | 27 (25%) | 46 (61%) | 46 (61%) |  |  |
| grade, post-pET |  |  |  |  |  | **1.6e-15** | **9.6e-13** |
| G1 | 36 (10%) | 23 (21%) | 0 | 13 (17%) | 0 |  |  |
| G2 | 214 (59%) | 84 (79%) | 46 (46%) | 62 (83%) | 22 (29%) |  |  |
| G3 | 114 (31%) | 0 | 61 (57%) | 0 | 53 (71%) |  |  |
| ER, baseline |  |  |  |  |  | **1** | **0.134** |
| > 10% | 358 (98%) | 106 (99%) | 107 (100%) | 74 (99%) | 71 (95%) |  |  |
| ≤ 10% | 4 (1%) | 1 (1%) | 0 | 0 | 3 (4%) |  |  |
| n.a. | 2 (1%) | 0 | 0 | 1 (1%) | 1 (1%) |  |  |
| ER, post-pET |  |  |  |  |  | **1** | **0.074** |
| > 10% | 358 (98%) | 106 (99%) | 107 (100%) | 75 (100%) | 70 (93%) |  |  |
| ≤ 10% | 6 (2%) | 1 (1%) | 0 | 0 | 5 (7%) |  |  |
| PR, baseline |  |  |  |  |  | **0.302** | **0.377** |
| > 10% | 305 (84%) | 101 (94%) | 96 (90%) | 57 (76%) | 51 (68%) |  |  |
| ≤ 10% | 59 (16%) | 6 (6%) | 11 (10%) | 18 (24%) | 24 (32%) |  |  |
| PR, post-pET |  |  |  |  |  | **1** | **0.201** |
| > 10% | 246 (68%) | 95 (89%) | 95 (89%) | 24 (32%) | 32 (43%) |  |  |
| ≤ 10% | 118 (32%) | 12 (11%) | 12 (11%) | 51 (68%) | 43 (57%) |  |  |
| HER2, baseline |  |  |  |  |  | **1<sup>d</sup>** | **0.502<sup>d</sup>** |
| IHC 0/1+ | 311 (85%) | 92 (86%) | 90 (84%) | 66 (88%) | 63 (84%) |  |  |
| IHC 2+, FISH-neg | 49 (13%) | 14 (13%) | 15 (14%) | 8 (11%) | 12 (16%) |  |  |
| IHC 2+, FISH-n.a. | 1 (0%) | 0 | 1 (1%) | 0 | 0 |  |  |
| IHC 2+, FISH-pos<sup>c</sup> | 3 (1%) | 1 (1%) | 1 (1%) | 1 (1%) | 0 |  |  |
| IHC 3+<sup>c</sup> | 0 | 0 | 0 | 0 | 0 |  |  |
| HER2, post-pET |  |  |  |  |  | **0.551<sup>d</sup>** | **0.144<sup>d</sup>** |
| IHC 0/1+ | 268 (74%) | 83 (78%) | 77 (72%) | 49 (65%) | 59 (79%) |  |  |
| IHC 2+, FISH-neg | 95 (26%) | 24 (22%) | 29 (27%) | 26 (35%) | 16 (21%) |  |  |
| IHC 2+, FISH-n.a. | 0 | 0 | 0 | 0 | 0 |  |  |
| IHC 2+, FISH-pos<sup>c</sup> | 0 | 0 | 0 | 0 | 0 |  |  |
| IHC 3+<sup>c</sup> | 1 (0%) | 0 | 1 (1%) | 0 | 0 |  |  |
| E-cadherin, baseline |  |  |  |  |  | 0.855<sup>d</sup> | 1<sup>d</sup> |
| pos. | 317 (87%) | 90 (84%) | 90 (84%) | 68 (91%) | 69 (92%) |  |  |
| neg. | 44 (12%) | 17 (16%) | 15 (14%) | 6 (8%) | 6 (8%) |  |  |
| n.a. | 3 (1%) | 0 | 2 (2%) | 1 (1%) | 0 |  |  |
| E-cadherin, post-pET |  |  |  |  |  | 0.855<sup>d</sup> | 0.724<sup>d</sup> |
| pos. | 314 (86%) | 90 (84%) | 89 (83%) | 67 (89%) | 68 (91%) |  |  |
| neg. | 44 (12%) | 17 (16%) | 15 (14%) | 5 (7%) | 7 (9%) |  |  |
| n.a. | 6 (2%) | 0 | 3 (3%) | 3 (4%) | 0 |  |  |
| Ki67, baseline |  |  |  |  |  | 0.495 | NA |
| 0% - 9% | 21 (6%) | 10 (9%) | 7 (7%) | 2 (3%) | 2 (3%) |  |  |
| 10% - 19% | 101 (28%) | 31 (29%) | 42 (39%) | 15 (20%) | 13 (17%) |  |  |
| 20% - 34% | 205 (56%) | 61 (57%) | 51 (47%) | 44 (58%) | 49 (65%) |  |  |
| 35% - 100% | 37 (10%) | 5 (5%) | 7 (7%) | 14 (19%) | 11 (15%) |  |  |
| Ki67, post-pET |  |  |  |  |  | **3.6e-05** | **2.2e-05** |
| 0% - 9% | 182 (50%) | 107 (100%) | 0 | 75 (100%) | 0 |  |  |
| 10% - 19% | 0 | 0 | 0 | 0 | 0 |  |  |
| 20% - 34% | 143 (39%) | 0 | 88 (82%) | 0 | 55 (73%) |  |  |
| 35% - 100% | 39 (11%) | 0 | 19 (18%) | 0 | 20 (27%) |  |  |
| Luminal Subtype<sup>e</sup>, baseline |  |  |  |  |  | 0.2 | 1 |
| LumA | 268 (74%) | 96 (90%) | 87 (81%) | 42 (57%) | 42 (56%) |  |  |
| LumB | 96 (26%) | 11 (10%) | 20 (19%) | 32 (43%) | 33 (44%) |  |  |
| Oncotype DX RS Group, baseline |  |  |  |  |  | 0.3 | NA |
| 1 (0 -11) | 78 (21%) | 30 (28%) | 30 (28%) | 9 (12%) | 9 (12%) |  |  |
| 2 (12 – 25) | 208 (57%) | 68 (64%) | 68 (64%) | 36 (48%) | 36 (48%) |  |  |
| 3 (26 – 100) | 78 (21%) | 9 (8%) | 9 (8%) | 30 (40%) | 30 (40%) |  |  |

Unless otherwise stated, the values are given in the format n (%), with n corresponding to the number of patients. The McNemar's chi-squared test for symmetry were used for statistical analysis between the matched pairs of Responder and Non-Responder. Significant differences are highlighted in bold. "NA" for the exact matched pairs therefore the p value is not available.
n.a. not available, ER estrogen receptor, PR progesterone receptor, pET preoperative endocrine therapy, RS recurrence score, TAM tamoxifen, AI aromatase inhibitors.
Case selection of the discovery cohort was done in a way that responders and non-responders were balanced for pT, pN, grade (baseline), histological type (baseline and post-pET), and RS group ("matching" to exclude confounding effects by dissimilar histological grades in R and NR).
a: Responder was defined as post-pET Ki67 < 10% and relative Ki67 decrease of ≥ 70% from baseline to post-pET.
b: Non-reponder was defined as post-pET Ki67 of ≥ 20% and relative Ki67 decrease of ≥ 20% from baseline to post-pET.
c: HER2-positivity in a minor subclone of <10% of tumor cells; not HER2-positive according to ASCO/CAP-Guidelines [76].
d: Comparison only between the first two category groups.
e: Luminal A: Ki67 baseline < 35% and PR baseline > 20%, Luminal B: Ki67 baseline ≥ 35% and PR baseline ≤ 20%

**Supplementary Table S1b.** Clinico-pathologic characteristics of the validation cohort.

|  | All | TAM |  | AI |  |  |  |
| --- | --- | --- | --- | --- | --- | --- | --- |
|  |  | Responder<sup>a</sup> | Non-Responder<sup>b</sup> | Responder<sup>a</sup> | Non-Responder<sup>b</sup> |  |  |
|  | n=270 | n=75 | n=80 | n=67 | n=48 | *P*<sub>TAM</sub> | *P*<sub>AI</sub> |
|  | n (%) | n (%) | n (%) | n (%) | n (%) |  |  |
| age |  |  |  |  |  | **2.4e-03** | 0.692 |
| < 50 | 97 (36%) | 35 (47%) | 59 (74%) | 2 (3%) | 1 (2%) |  |  |
| 50 – 59 | 83 (31%) | 34 (45%) | 18 (22%) | 16 (24%) | 15 (31%) |  |  |
| ≥ 60 | 90 (33%) | 6 (8%) | 3 (4%) | 49 (73%) | 32 (67%) |  |  |
| histology, baseline |  |  |  |  |  | **3.0e-07** | 0.666 |
| NST | 222 (82%) | 48 (64%) | 77 (96%) | 57 (85%) | 40 (85%) |  |  |
| ILBC | 41 (15%) | 22 (29%) | 2 (3%) | 10 (15%) | 7 (15%) |  |  |
| Others<sup>c</sup> | 7 (3%) | 5 (7%) | 1 (1%) | 0 | 1 (2%) |  |  |
| pT stage |  |  |  |  |  | **0.017** | 0.816 |
| pT1 | 145 (54%) | 43 (57%) | 39 (49%) | 38 (57%) | 25 (52%) |  |  |
| pT2 | 113 (42%) | 27 (36%) | 40 (50%) | 26 (39%) | 20 (42%) |  |  |
| pT3 | 8 (3%) | 5 (7%) | 0 | 2 (3%) | 1 (2%) |  |  |
| pT4 | 4 (1%) | 0 | 1 (1%) | 1 (1%) | 2 (4%) |  |  |
| pN stage |  |  |  |  |  | 0.390 | 0.206 |
| pN0 | 226 (84%) | 60 (80%) | 69 (86%) | 59 (88%) | 38 (79%) |  |  |
| pN1+ | 44 (16%) | 15 (20%) | 11 (14%) | 8 (12%) | 10 (21%) |  |  |
| grade, baseline |  |  |  |  |  | **7.5e-04** | **1.7e-03** |
| G1 | 22 (8%) | 14 (19%) | 4 (5%) | 3 (4%) | 0 |  |  |
| G2 | 143 (53%) | 51 (68%) | 48 (60%) | 33 (49%) | 11 (23%) |  |  |
| G3 | 106 (39%) | 10 (13%) | 28 (35%) | 31 (47%) | 37 (77%) |  |  |
| grade, post-pET |  |  |  |  |  | **5.2e-23** | **3.2e-17** |
| G1 | 33 (12%) | 16 (21%) | 0 | 17 (25%) | 0 |  |  |
| G2 | 146 (54%) | 58 (77%) | 24 (30%) | 49 (73%) | 15 (31%) |  |  |
| G3 | 91 (34%) | 1 (1%) | 56 (70%) | 1 (2%) | 33 (69%) |  |  |
| ER, baseline |  |  |  |  |  | 0.497 | 1 |
| > 10 | 266 (99%) | 75 (100%) | 78 (98%) | 66 (99%) | 47 (98%) |  |  |
| ≤ 10 | 4 (1%) | 0 | 2 (2%) | 1 (1%) | 1 (2%) |  |  |
| ER, post-pET |  |  |  |  |  | 0.611 | 1 |
| > 10 | 266 (99%) | 73 (97%) | 79 (99%) | 66 (99%) | 48 (100%) |  |  |
| ≤ 10 | 4 (1%) | 2 (3%) | 1 (1%) | 1 (1%) | 0 |  |  |
| PR, baseline |  |  |  |  |  | 0.133 | **0.012** |
| > 10 | 227 (84%) | 72 (96%) | 71 (89%) | 55 (82%) | 29 (60%) |  |  |
| ≤ 10 | 43 (12%) | 3 (4%) | 9 (11%) | 12 (18%) | 19 (40%) |  |  |
| PR, post-pET |  |  |  |  |  |  |  |
| > 10 | 167 (62%) | 67 (89%) | 69 (86%) | 23 (34%) | 8 (17%) | 0.629 | 0.054 |
| ≤ 10 | 103 (38%) | 8 (11%) | 11 (14%) | 44 (66%) | 40 (83%) |  |  |
| HER2, baseline |  |  |  |  |  |  |  |
| IHC 0/1+ | 242 (90%) | 68 (91%) | 72 (90%) | 59 (88%) | 43 (90%) | 1<sup>e</sup> | 1<sup>e</sup> |
| IHC 2+, FISH-neg | 28 (10%) | 7 (9%) | 8 (10%) | 8 (12%) | 5 (10%) |  |  |
| IHC 2+, FISH-n.a. | 0 | 0 | 0 | 0 | 0 |  |  |
| IHC 2+, FISH-pos<sup>d</sup> | 0 | 0 | 0 | 0 | 0 |  |  |
| IHC 3+<sup>d</sup> | 0 | 0 | 0 | 0 | 0 |  |  |
| HER2, post-pET |  |  |  |  |  | 0.573<sup>e</sup> | 0.371<sup>e</sup> |
| IHC 0/1+ | 208 (77%) | 56 (75%) | 63 (79%) | 54 (81%) | 35 (73%) |  |  |
| IHC 2+, FISH-neg | 62 (23%) | 19 (25%) | 17 (21%) | 13 (19%) | 13 (27%) |  |  |
| IHC 2+, FISH-n.a. | 0 | 0 | 0 | 0 | 0 |  |  |
| IHC 2+, FISH-pos<sup>d</sup> | 0 | 0 | 0 | 0 | 0 |  |  |
| IHC 3+<sup>d</sup> | 0 | 0 | 0 | 0 | 0 |  |  |
| E-cadherin, baseline |  |  |  |  |  | **1.2e-05<sup>e</sup>** | 0.761<sup>e</sup> |
| pos. | 218 (81%) | 50 (67%) | 73 (94%) | 55 (82%) | 38 (79%) |  |  |
| neg. | 44 (16%) | 22 (29%) | 6 (4%) | 11 (16%) | 8 (17%) |  |  |
| n.a. | 8 (3%) | 3 (4%) | 1 (2%) | 1 (2%) | 2 (4%) |  |  |
| E-cadherin, post-pET |  |  |  |  |  | **1.7e-04<sup>e</sup>** | 0.516<sup>e</sup> |
| pos. | 218 (81%) | 50 (67%) | 73 (91%) | 53 (79%) | 42 (88%) |  |  |
| neg. | 49 (18%) | 24 (32%) | 6 (8%) | 13 (19%) | 6 (12%) |  |  |
| n.a. | 3 (1%) | 1 (1%) | 1 (1%) | 1 (2%) | 0 |  |  |
| Ki67, baseline |  |  |  |  |  | **1.2e-07** | **2.1e-05** |
| 0-9 | 22 (8%) | 12 (16%) | 0 | 10 (15%) | 0 |  |  |
| 10-19 | 103 (38%) | 52 (69%) | 36 (45%) | 13 (19%) | 2 (4%) |  |  |
| 20-34 | 83 (31%) | 10 (13%) | 35 (44%) | 25 (37%) | 12 (25%) |  |  |
| 35-100 | 63 (23%) | 1 (1%) | 9 (11%) | 19 (28%) | 34 (71%) |  |  |
| Ki67, post-pET |  |  |  |  |  | **NA** | **NA** |
| 0-9 | 142 (53%) | 75 (100%) | 0 | 67 (100%) | 0 |  |  |
| 10-19 | 0 | 0 | 0 | 0 | 0 |  |  |
| 20-34 | 87 (32%) | 0 | 46 (58%) | 0 | 41 (85%) |  |  |
| 35-100 | 41 (15%) | 0 | 34 (42%) | 0 | 7 (15%) |  |  |
| Luminal Subtype<sup>f</sup>, baseline |  |  |  |  |  | **6.2e-03** | **2.1e-06** |
| LumA | 175 (65%) | 70 (93%) | 62 (78%) | 37 (55%) | 6 (13%) |  |  |
| LumB | 95 (35%) | 5 (7%) | 18 (22%) | 30 (44%) | 42 (87%) |  |  |
| Oncotype DX RS Group, baseline |  |  |  |  |  | **1.9e-08** | **2.1e-12** |
| 1 (0 -11) | 57 (21%) | 31 (41%) | 5 (6%) | 20 (30%) | 1 (2%) |  |  |
| 2 (12 – 25) | 156 (58%) | 41 (55%) | 56 (70%) | 43 (64%) | 16 (33%) |  |  |
| 3 (26 – 100) | 57 (21%) | 3 (4%) | 19 (24%) | 4 (6%) | 31 (65%) |  |  |

Unless otherwise stated, the values are given in the format n (%), with n corresponding to the number of patients. For the statistical analysis between the Responder and Non-Responder groups, Fisher's exact test was used. Significant differences are highlighted in bold. "NA" indicates data with clear separation therefore the p value is not available.
n.a. not available, ER estrogen receptor, PR progesterone receptor, pET preoperative endocrine therapy, RS recurrence score, TAM tamoxifen, AI aromatase inhibitors.
Case selection criteria in the validation cohort were made less strict and no matching between NR and R.
a: Responder was defined as post-pET Ki67 <10%.
b: Non-reponder was defined as post-pET Ki67 of ≥ 20%.
c: Others: Tubular, Medullary, Mucinous.
d: HER2-positivity in a subclone <10% of tumor cells. not HER2-positive according to ASCO/CAP-Guidelines [76]
e: Comparison only between the first two category groups.
f: Luminal A: Ki67 baseline < 35% and PR baseline > 20%, Luminal B: Ki67 baseline ≥ 35% and PR baseline ≤ 20%

**Supplementary Table S2.** Clinico-pathologic features of the TCGA-BRCA sub-cohort.

|  | All | TAM-like |  | AI-like |  |  |  |
| --- | --- | --- | --- | --- | --- | --- | --- |
|  |  | PERCI<br>TAM 450k<br>*low* | PERCI<br>TAM 450k<br>*high* | PERCI<br>AI 450k<br>*low* | PERCI<br>AI 450k<br>*high* |  |  |
|  | **n=269** | **n=26** | **n=49** | **n=143** | **n=51** | *P<sub>TAM like</sub>* | *P<sub>AI like</sub>* |
|  | n (%) | n (%) | n (%) | n (%) | n (%) |  |  |
| Age |  |  |  |  |  | **0.22** | **1.9e-03** |
| < 50 | 73 (26%) | 19 (73%) | 42 (86%) | 6 (4%) | 6 (12%) |  |  |
| 50 - 59 | 78 (30%) | 7 (27%) | 7 (14%) | 40 (28%) | 24 (47%) |  |  |
| ≥ 60 | 118 (44%) | 0 | 0 | 97 (68%) | 21 (41%) |  |  |
| Menopause |  |  |  |  |  | **1** | **1** |
| Pre | 75 (28%) | 26 (100%) | 49 (100%) | 0 | 0 |  |  |
| Post | 194 (72%) | 0 | 0 | 143 (100%) | 51 (100%) |  |  |
| Histology |  |  |  |  |  | **0.032** | **0.287** |
| NST | 190 (71%) | 15 (58%) | 40 (82%) | 96 (67%) | 39 (76%) |  |  |
| ILBC | 79 (29%) | 11 (42%) | 9 (18%) | 47 (33%) | 12 (24%) |  |  |
| pT Stage |  |  |  |  |  | **0.458** | **0.026** |
| pT1 | 77 (29%) | 3 (12%) | 10 (21%) | 55 (38%) | 9 (18%) |  |  |
| pT2 | 156 (58%) | 17 (65%) | 32 (65%) | 72 (50%) | 35 (68%) |  |  |
| pT3 | 35 (13%) | 6 (23%) | 7 (24%) | 15 (11%) | 7 (14%) |  |  |
| pT4 | 1 (0%) | 0 | 0 | 1 (1%) | 0 |  |  |
| pN Stage |  |  |  |  |  | **1** | **1** |
| pN0 | 126 (47%) | 12 (46%) | 17 (35%) | 71 (50%) | 26 (51%) |  |  |
| pN1+ | 143 (53%) | 14 (54%) | 32 (65%) | 72 (50%) | 25 (49%) |  |  |
| Grade |  |  |  |  |  | **0.314** | **0.026** |
| G1 | 74 (27%) | 8 (31%) | 10 (21%) | 49 (34%) | 7 (14%) |  |  |
| G2 | 123 (46%) | 15 (58%) | 24 (49%) | 59 (41%) | 25 (49%) |  |  |
| G3 | 57 (21%) | 3 (11%) | 13 (26%) | 27 (19%) | 14 (28%) |  |  |
| NA | 15 (6%) | 0 | 2 (4%) | 8 (6%) | 5 (10%) |  |  |
| ER |  |  |  |  |  | **1** | **1** |
| Positive | 269 (100%) | 26 (100%) | 49 (100%) | 143 (100%) | 51 (100%) |  |  |
| Negative | 0 | 0 | 0 | 0 | 0 |  |  |
| PR |  |  |  |  |  | **1** | **0.528** |
| Positive | 228 (85%) | 22 (85%) | 42 (86%) | 123 (86%) | 41 (80%) |  |  |
| Negative | 40 (15%) | 4 (15%) | 7 (14%) | 19 (13%) | 10 (20%) |  |  |
| Indeterminate | 1 (0%) | 0 | 0 | 1 (1%) | 0 |  |  |
| HER2 |  |  |  |  |  | **0.421<sup>b</sup>** | **0.107<sup>b</sup>** |
| Negative | 193 (72%) | 17 (65%) | 37 (76%) | 107 (75%) | 32 (63%) |  |  |
| Equivocal<sup>a</sup> | 76 (28%) | 9 (35%) | 12 (24%) | 36 (25%) | 19 (37%) |  |  |
| Positive | 0 | 0 | 0 | 0 | 0 |  |  |
| Progression-free survival (PFS) status<sup>c</sup> |  |  |  |  |  | **0.012** | **0.084** |
| 0 | 247 (92%) | 26 (100%) | 39 (80%) | 137 (96%) | 45 (88%) |  |  |
| 1 | 22 (8%) | 0 | 10 (20%) | 6 (4%) | 6 (12%) |  |  |

Unless otherwise stated, the values are given in the format n (%), with n corresponding to the number of patients. For statistical analysis between the PERCI TAM/AI low and PERCI TAM/AI high, Fisher's exact test (FET) was used. Significant differences are highlighted in bold.
ER estrogen receptor, PR progesterone receptor, PFS progression-free survival.
a: Her2 equivocal was defined in TCGA either with scores of 2+ in the immunohistochemistry (IHC) assay or with a HER2/CEP17 ratio between 1.8 and 2.2 in Fluorescence in situ hybridization (FISH).
b: Comparison only between the first two category groups.
c: 0 censored, 1 progression

